# Estimating the burden of SARS-CoV-2 in France

**DOI:** 10.1101/2020.04.20.20072413

**Authors:** Henrik Salje, Cécile Tran Kiem, Noémie Lefrancq, Noémie Courtejoie, Paolo Bosetti, Juliette Paireau, Alessio Andronico, Nathanaël Hozé, Jehanne Richet, Claire-Lise Dubost, Yann Le Strat, Justin Lessler, Daniel Levy Bruhl, Arnaud Fontanet, Lulla Opatowski, Pierre-Yves Boelle, Simon Cauchemez

## Abstract

France has been heavily affected by the SARS-CoV-2 epidemic and went into lockdown on the 17th March 2020. Using models applied to hospital and death data, we estimate the impact of the lockdown and current population immunity. We find 2.6% of infected individuals are hospitalized and 0.53% die, ranging from 0.001% in those <20y to 8.3% in those >80y. Across all ages, men are more likely to be hospitalized, enter intensive care, and die than women. The lockdown reduced the reproductive number from 3.3 to 0.5 (84% reduction). By 11 May, when interventions are scheduled to be eased, we project 3.7 million (range: 2.3-6.7) people, 5.7% of the population, will have been infected. Population immunity appears insufficient to avoid a second wave if all control measures are released at the end of the lockdown.

## Main

The worldwide pandemic of SARS-CoV-2, the coronavirus which causes COVID-19, has resulted in unprecedented responses, with many affected nations confining residents to their homes. Much like the rest of Europe, France has been hit hard by the epidemic and went into lockdown on the 17th March. It was hoped that this would result in a sharp decline in ongoing spread, as was observed when China locked down following the initial emergence of the virus (*1*). Following a reduction in cases, the government has announced it will ease restrictions on the 11th May. In order to appropriately exit from the lockdown, we need to understand the underlying level of population immunity and infection, identify those most at risk for severe disease and the impact of current control efforts.

Daily reported numbers of hospitalizations and deaths only provide limited insight into the state of the epidemic. Many people will either develop no symptoms or symptoms so mild they will not be detected through healthcare-based surveillance. The concentration of hospitalized cases in older individuals has led to hypotheses that there may be widespread ‘silent’ transmission in younger individuals. For example, it has been suggested that up to half the UK population may have been infected by the middle of March, despite under 100 deaths reported at that time (*2*). If the majority of the population is infected, viral transmission would slow, potentially reducing the need for the stringent intervention measures currently employed.

Here, we present a suite of modelling analyses to characterize the dynamics of SARS-CoV-2 transmission in France and the impact of the lockdown on these dynamics. We elucidate the risk of SARS-CoV-2 infection and severe outcomes by age and sex and estimate the current proportion of the national and regional populations that have been infected and might be at least temporarily immune (*3*). These models support healthcare planning of the French government by forecasting Intensive Care Unit (ICU) bed capacity requirements.

As of 14 April 2020, there had been 71,903 incident hospitalizations due to SARS-CoV-2 reported in France and 10,129 deaths in hospitals, with the east of the country and the capital, Paris, particularly affected (Figure 1A-B). The mean age of hospitalized patients was 68y and the mean age of the deceased was 79y with 50.0% of hospitalizations occurring in individuals >70y and 81.6% of deaths within that age bracket; 56.2% of hospitalizations and 60.3% of deaths were male (Figures 1C-E). Hospitalization and death data only capture the most severe infections. To reconstruct the dynamics of all infections, including mild ones, we jointly analyze them with data documenting the risk of death among persons infected by SARS-CoV-2 coming from a detailed investigation of an outbreak aboard the Princess Diamond cruise ship where all passengers were subsequently tested (719 infections and 13 deaths). By coupling the French passive surveillance hospital data with the active surveillance performed aboard the Princess Diamond, we can disentangle the risk of being hospitalized in those infected d from the underlying probability of infection (*4*–*6*).

**Figure 1.**
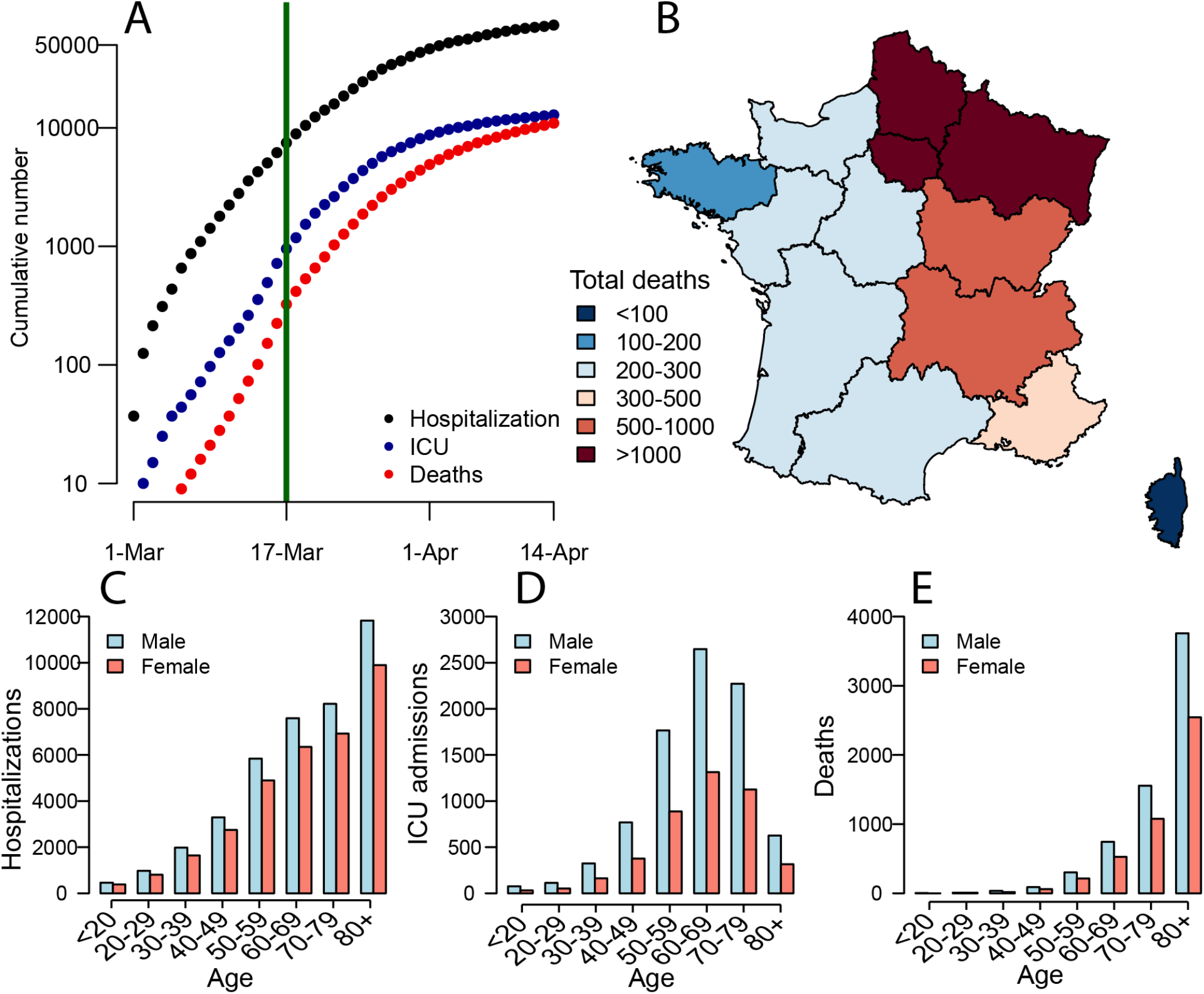
**(A)** Cumulative number of hospitalizations, ICU admissions and deaths from SARS-CoV-2 in France. The green line indicates the time when the lockdown was put in place in France. **(B)** Distribution of deaths in France. Number of **(C)** hospitalizations, **(D)** ICU and **(E)** deaths by age group and sex in France.

We find that 2.6% of infected individuals are hospitalized (95% CrI: 1.4-4.4), ranging from 0.09% (95% CrI: 0.05-0.2) in females under <20y to 31.4% (95% CrI: 16.7-52.6) in males >80y (Figure 2A, Table S1). Once hospitalized, 18.2% (95% CrI: 18.0%-18.6%) patients enter ICU after a mean delay of 1.5 days (Figure S1). There is an increasing probability of entering ICU with age - however, this drops for those >70y (Figure 2B, Table S2). Overall, 20.0% (95% CrI: 19.6-20.4) of hospitalized individuals go on to die (Figure 2C). The overall probability of death among those infected (the Infection Fatality Ratio, IFR) is 0.53% (95% CrI: 0.28-0.88), ranging from 0.001% in those under 20y to 8.3% (95% CrI: 4.4-13.9) in those >80y (Figure 2D, Table S2). Our estimate of overall IFR is similar to other recent studies that found values of between 0.5%-0.7% for the Chinese epidemic (*5, 8, 9*). We find men have a consistently higher risk than women of hospitalization (RR 1.26, 95% CrI: 1.21-1.31), ICU admission once hospitalized (RR: 1.69, 95% CrI: 1.61-1.78) and death (RR: 1.45, 95% CrI: 1.26-1.74) across all age groups (Figure S2).

**Figure 2.**
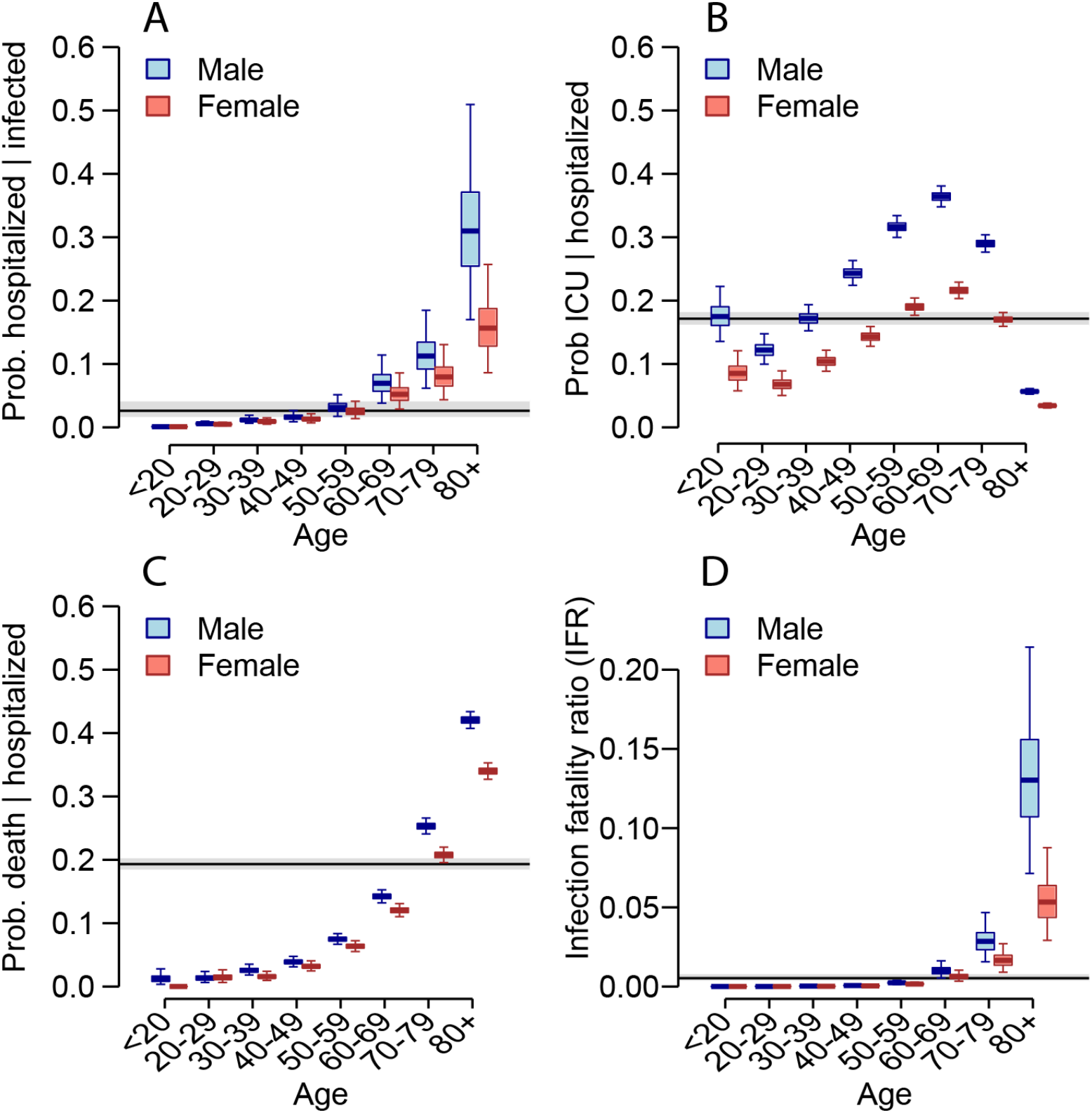
**(A)** Probability of hospitalization among those infected as a function of age and sex. **(B)** Probability of ICU admission among those hospitalized as a function of age and sex. **(C)** Probability of death among those hospitalized as a function of age and sex. **(D)** Probability of death among those infected as a function of age and sex. For each panel, the black line and grey shaded region represents the overall mean across all ages. The boxplots represent the 2.5, 25, 50, 75 and 97.5 percentiles of the posterior distributions.

We identify two clear subpopulations in delays between hospitalization and death: individuals that die quickly upon hospital admission (15% of fatal cases, mean time to death of 0.67 days) and individuals who die after longer time periods (85% of fatal cases, mean time to death of 13.2 days) (Figure S3). The proportion of fatal cases who die rapidly remains approximately constant across age-groups (Figure S4, Table S3). These observations, combined with the substantial differences in risk of death by age and sex, suggest complex patterns of disease manifestation. The underlying mechanisms that can explain why subsets of the population are substantially more at risk than others remain unclear although a role for immunopathogenesis has been proposed, including through antibody-dependent enhancement, where non-neutralizing antibodies trigger an immune response through cytokine storms, and cell-based enhancement, such as with allergic inflammation (*10*–*12*). Understanding these processes with regards to SARS-CoV-2 will be key to the development of a safe vaccine.

We next fit national and regional transmission models for the epidemic to ICU admission and bed occupancy (Figure 3A-B, Figure S5, Tables S4-S6). We find that the basic reproductive number R0 prior to the implementation of the lockdown was 3.31 (95% CrI: 3.18-3.43). At a national level, the lockdown resulted in a 84% reduction in transmission, with the reproduction number R dropping to 0.52 (95% CrI: 0.50-0.55). We forecast that by the 11th May, 3.7 million (range 2.3 - 6.7, when accounting for uncertainty in the probability of entering ICU) people will have been infected, representing 5.7% (range 3.5 - 10.3) of the French population (Figure 3E). This proportion will be 12.3% (range 7.9-21.3) in Ile-de-France, which includes Paris, and 11.8% (range 7.4-20.5) in Grand Est, the two most affected regions of the country (Figure 3D, Figure S5). Assuming a basic reproductive number of R0=3.3, it would require around 70% of the population to be immune for the epidemic to be controlled by immunity alone. Our results therefore strongly suggest that, without a vaccine, herd immunity on its own will be insufficient to avoid a second wave at the end of the lockdown. Efficient control measures need to be maintained beyond the 11th May.

**Figure 3.**
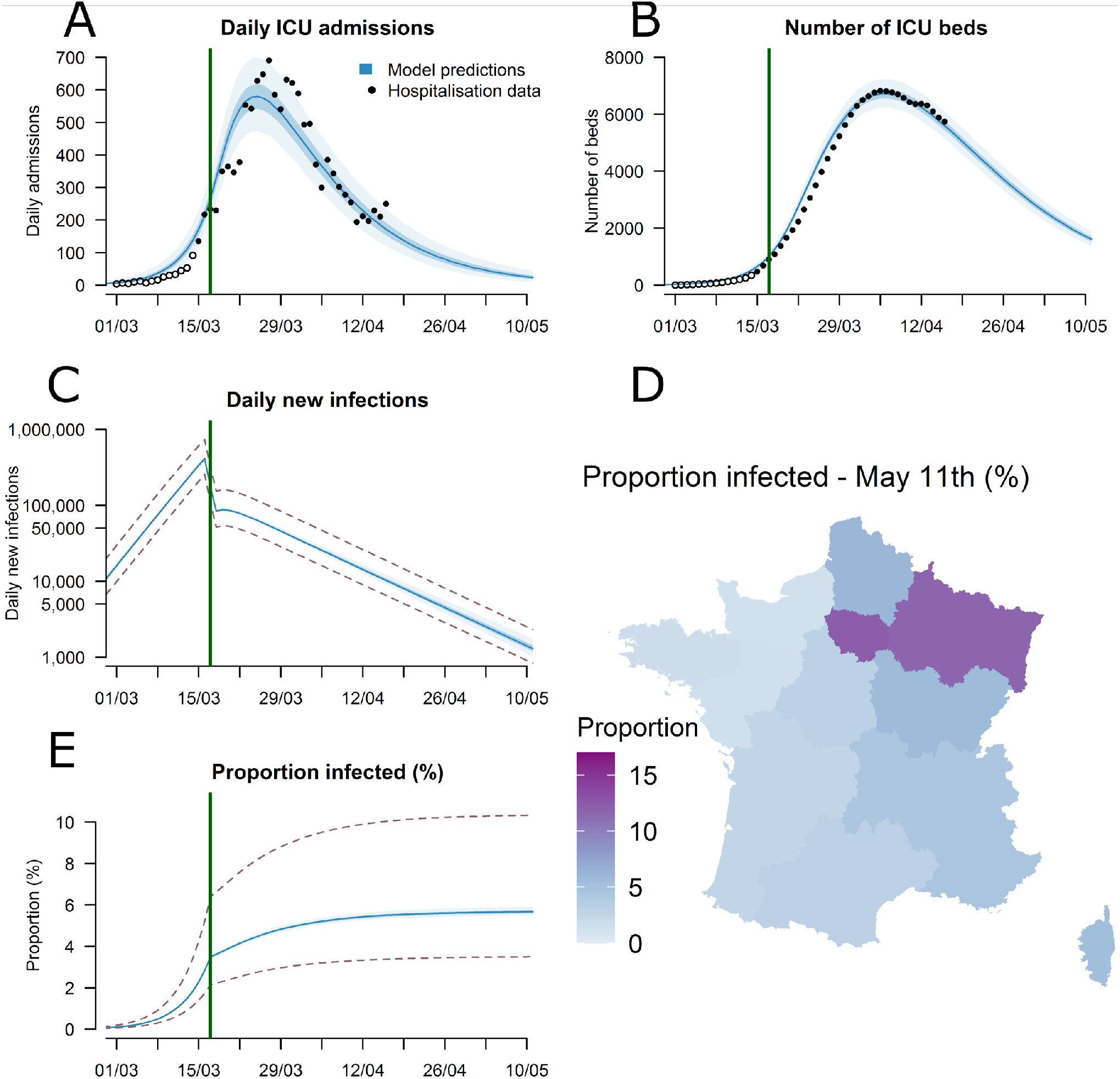
**(A)** Daily admissions in ICU in metropolitan France. **(B)** Number of ICU beds occupied in metropolitan France. **(C)** Daily new infections in metropolitan France (logarithmic scale). **(D)** Predicted proportion of the population infected by May 11th 2020 for each of the 13 regions in metropolitan France. **(E)** Predicted proportion of the population infected in metropolitan France. The black circles in panels A and B represent hospitalization data used for the calibration and the open circles hospitalization data that were not used for calibration. The dotted lines in panels C and E represent the 95% uncertainty range stemming from the uncertainty in the probability of entering ICU following infection.

Our model projections can help inform the ongoing and future response to COVID-19. National ICU daily admissions have gone from 700 at the end of March to 220 on the 14th April. If current trends continue, by the 11th May we project between 10 and 45 ICU daily admissions, between 1370 and 1900 ICU beds occupied by COVID-19 cases as well as 1300 (range 840 - 2300) daily infections, down from between 270,000-770,000 immediately prior to the lockdown. However, it is important to emphasize that these dynamics and forecasts may change quickly and so extreme caution is needed. For example, after an initial important drop, we note that ICU admissions have plateaued in the last few days. In addition, other external factors, such as temperature fluctuations may affect the dynamics over time (*13*).

Using our framework, we are able to recover the observed number of hospitalizations by age and sex in France and the number of deaths in the Princess Diamond (Figure S6). As a validation, our approach is also able to recover parameters in simulated datasets where the true values are known (Figure S7). We run a suite of sensitivity analyses that considers additional future COVID-19 deaths on the Princess Diamond, longer delays between symptom onset and ICU admission, equal attack rates across all ages, reduced infectivity in younger individuals, a contact matrix with unchanged structure before/during the lockdown and one with extremely high isolation of elderly individuals during the lockdown. These different scenarios result in mean IFRs ranging from 0.4% to 0.7%, the level of immunity in the population by the 11 May ranging from 1.9-12.5, the number of daily infections at this date ranging from 200-4600 and a range of reproductive numbers post lockdown of 0.38-0.64 (Figure 4, Figures S8-S10).

**Figure 4.**
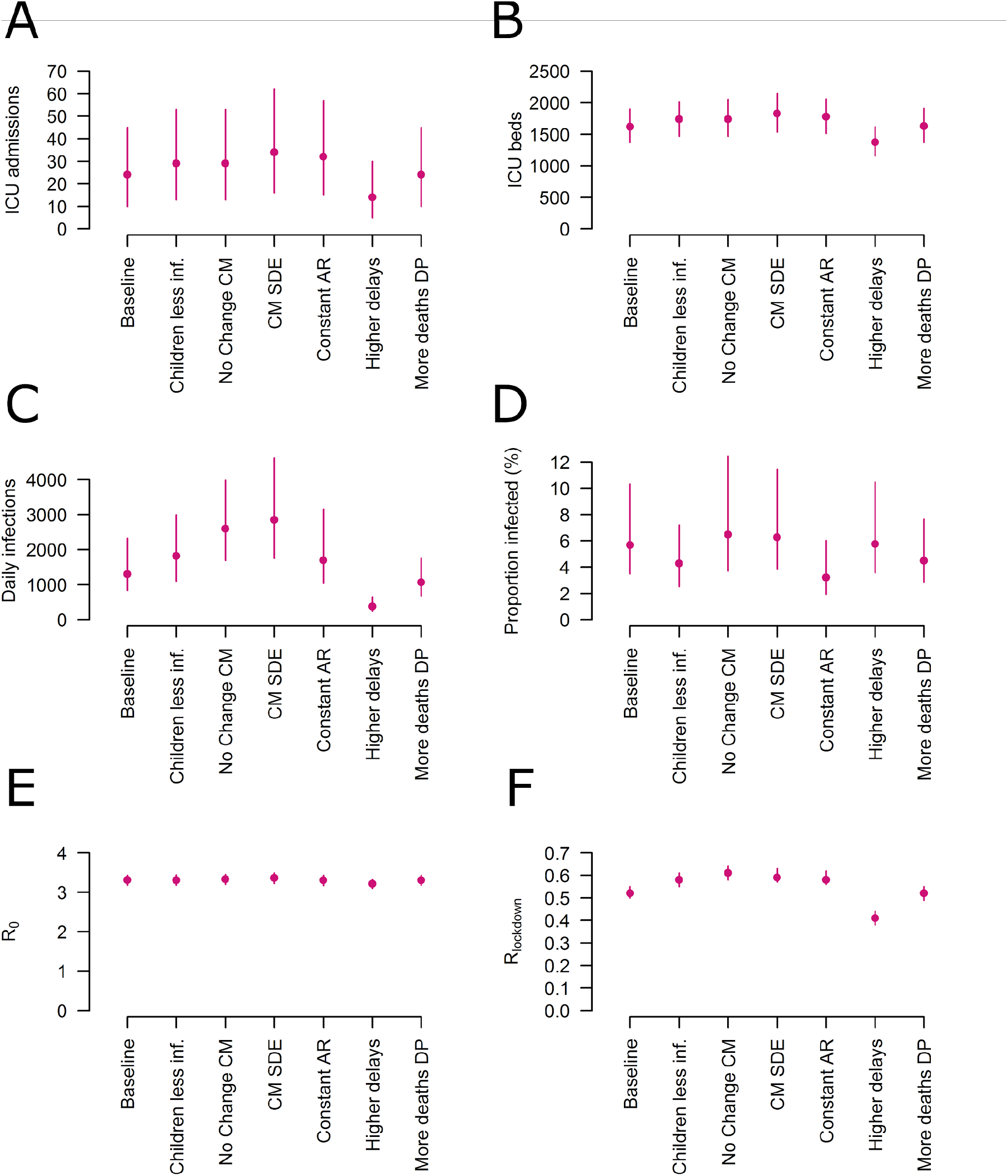
Sensitivity analysis considering different modelling assumptions. **(A)** Predicted daily ICU admissions on May 11th. **(B)** Predicted ICU beds on May 11th. **(C)** Predicted daily new infections on May 11th. **(D)** Predicted proportion of the population infected by May 11th. **(E)**Estimated basic reproduction number before lockdown. **(F)** Estimated reproduction number during lockdown. The different scenarios correspond to: Children less inf. - Individuals <20y are half as infectious as adults; No Change CM - the structure of the contact matrix is modified by the lockdown; CM SDE - Contact matrix after lockdown with social distancing of the elderly; Constant AR - Attack rates are constant across age groups; Higher delays - 9 days on average between illness onset and ICU admission instead of 7 days; More deaths DP - Three additional deaths will occur amongst the six passengers of the Diamond Princess cruise ship that are still in ICU. For estimates of ICU admissions, ICU beds and reproduction numbers before and after lockdown, we report 95% credible intervals. For estimates of daily new infections and proportion of the population infected by May 11th, we report the 95% uncertainty range stemming from the uncertainty in the probability of entering ICU given infection.

Here, we focused on deaths occurring in hospitals. There are also non-hospitalized COVID-19 deaths. For example there have been >6,000 non-hospitalized deaths in retirement homes in France (*14*). We explicitly removed these communities from our analyses as transmission dynamics may be different in these closed communities, therefore these do not affect our estimates of immunity in the general population. A number of additional non-hospitalized deaths may also be occurring, in which case we would underestimate the proportion infected.

This study shows the massive impact the French lockdown had on SARS-CoV-2 transmission. It estimates underlying probabilities of infection, hospitalization and death, which is essential for the interpretation of COVID-19 surveillance data. The forecasts we provide can inform planning of ICU bed occupancy and lockdown exit strategies. The estimated low level of immunity against SARS-CoV-2 indicates that efficient control measures that limit transmission risk will have to be maintained beyond the 11th May to avoid a rebound of the epidemic.

## Methods

### Case data

We work with daily hospitalization and death data from the SI-VIC database, maintained by the ANS (Agence du Numérique en Santé, formerly named ASIP) and sent daily to Santé Publique France, the French national public health agency. This database provides real time data on the COVID-19 patients hospitalized in French public and private hospitals, including their age, date of hospitalization and region. All cases are either biologically confirmed or present with a computed tomographic image highly suggestive of SARS-CoV-2 infection. The SI-VIC web portal was activated for the COVID-19 epidemic on 13 March 2020, with a progressive increase in the number of hospitals transmitting data. Every day, we receive a case line-list with the latest hospitalization status of each patient. We collate the daily files for metropolitan France to reconstruct individual trajectories from hospital admission to discharge or death. We report as hospitalized in ICU patients hospitalized in “*Hospitalisation réanimatoire (Réanimation, soins intensifs or unité de surveillance continue)*”.

Since the status of each patient is updated with a few days of delay, we correct the observed time series for these reporting delays. Let *H*_*t,T*_denote the number of hospital admissions that were reported for time *t* at time *T* of the epidemic (*t*≤*T*). Let *p*_*t,T*_ denote the probability that a hospital admission that occured at time *t*has been reported before time *T*. We estimate this probability from the cumulative distribution of hospital admissions reporting delays estimated from the SI-VIC database. We correct the observed time series of hospital admissions at time *T* by sampling the expected number of hospital admissions at time *t <u>H</u>*_*t,T*_ that have not been reported yet from:

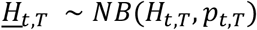

where *NB*is a negative binomial distribution. We then compute the expected number of hospital admissions corrected for reporting delays as:

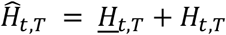

In order to take into account the variations of the reporting delays with the day of the week (from e.g., reduced reporting over weekends), we estimate different probabilities *p*_*t,T*_ according to the day of the week of *t* and *T*. We apply the same method to correct the daily time series of ICU admissions, deaths and discharges, as well as ICU releases in order to compute the corrected times series of occupied ICU beds.

Since the SI-VIC web portal started recording COVID-19 cases on March 13^th^, a certain number of hospital admissions prior to or around this date may have been missed. We therefore adjusted the data from this database before March 15^th^ by using data on hospital admissions collected by the OSCOUR® network. This surveillance system was created in 2004. It collects data on patients presenting to emergency departments in all regions of France. To correct the SI-VIC data we computed the median ratio between hospital admissions in the SI-VIC and in the OSCOUR® datasets past March 15^th^ and multiplied by the same ratio to the OSCOUR® data before March 15^th^ (see Figure S11).

### Active surveillance data

The Princess Diamond is a cruise ship that suffered a SARS-CoV-2 outbreak in early February 2020. All individuals on board were tested. Out of 3711 passengers, 712 tested positive (*15, 16*). The age distribution of the positive individuals is available for a subset of 619 individuals (*15*). We assume that the age distribution of the remaining 93 individuals who tested positive is the same. There have been 13 deaths, seven were individuals in their 70s, four were in their 80s, one in their 60s. No age was reported for one death.

### Estimating delays from hospitalization to death and from hospitalization to ICU

In a growing epidemic, the time to death among individuals who have already experienced the outcome will be an underestimate of overall time to death, as many of those who take longer will not yet have died. We need to account for these delays when estimating the infection fatality ratio as otherwise we would underestimate the probability of death (*8*).

To capture the delay to death for the different age groups we use data from cases throughout France that had dates of hospitalization and dates of death. We assume that the delays follow a lognormal distribution as this has previously been shown to work well for SARS-CoV-2 infections (*17*).

We use the number of hospitalizations on a given day to account for the state of the epidemic at that time, similar to what has previously been used (*5*). We note that a subset of individuals die within a short period of time after entering hospital. We therefore use a mixture distribution composed of an exponential distribution for those that die within a short delay and a lognormal distribution for those that die after longer delays (Figure S3).

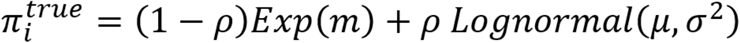

We denote by *π*_*i*_^*true*^ the true probability density function (pdf) of the delay, and *π*_*i*_^*obs*^ the observed density, which will be biased to be right skewed as most individuals will not have had their outcome. We denote by *π*_*i*_^*true*^ and *π*_*i*_^*obs*^ their cumulative density functions (cdf), respectively. We can approximate the expected delay distribution *π*_*i*_^*exp*^, for a given age group i, at a given time T during the epidemic, thereby adjusting for the stage of the epidemic, given the true pdf for the delay *π*_*i*_^*true*^ using the following adjustment:

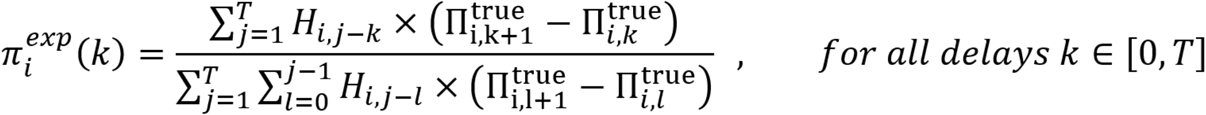

where *H*_*i,j*_ is the total number of hospitalized cases of age i at time j.

For the correct pdf *π*_*i*_^*true*^, we should have:

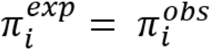

We estimate parameters of the true delay from hospitalization to death distribution *π*_*i*_^*true*^ for each group in turn by minimizing the sum of squared error (SSE) of the distribution *π*_*i*_^*exp*^ to the observed data *π*_*i*_^*obs*^. Given the small number of deaths in younger age groups, we consider three age groups: <70y, 70-80, 80+. To get an overall estimate, we also repeat the calculation using all individuals across all age groups.

To fit the delays from hospitalization to ICU admission we use the same approach, however, we consider the delays are constant across age groups and that they follow an exponential distribution (Figure S1).

### Modeling the risk of hospitalization, ICU admission and death

We consider the population of mainland France for the transmission model in eight age bands (<20y, 20-29, 30-39, 40-49, 50-59, 60-69, 70-79, 80+) and consider males and females separately. We exclude the population in retirement communities (N=730,000 individuals, mainly over the age of 70 and 74% female), as there have been a number of outbreaks in these enclosed communities and the underlying risk of infection in these locations is unlikely to be the same as the wider population. Deaths in these communities are not captured in hospital records.

Outside retirement communities, we assume that all recorded deaths occurred in hospital and that the probability of death is linked to age and sex.

As SARS-CoV-2 is principally transmitted from close contact between individuals, we assume that the probability of infection is proportional to the number of contacts an individual makes, which has previously been measured for individuals in France (*7*).

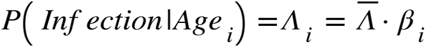

Where *<u>Λ</u>* represents the mean cumulative probability of having been infected across the entire population and *β*_*i*_ represents the relative risk of infection for an individual of age *i* compared to a randomly selected person from the population. For *β*_*i*_, we use the mean number of contacts that an individual of age group *i* has on a daily basis as measured in France, weighted by the proportion of the population that is within age group *i (7)*.

In order to disentangle the underlying probability of infection from the probability of hospitalization and death, we use the results of an active surveillance campaign in a different population (cruise ship) where all individuals were tested, and therefore the probability of detection is not linked to the presence of severe disease that requires hospitalization. Where the age of the individuals is reported, we can estimate the probability of death given infection for each age group.

For the passive French hospital surveillance system, we use a Poisson Likelihood for the number of hospitalizations, ICU admissions and deaths within each age group and sex.

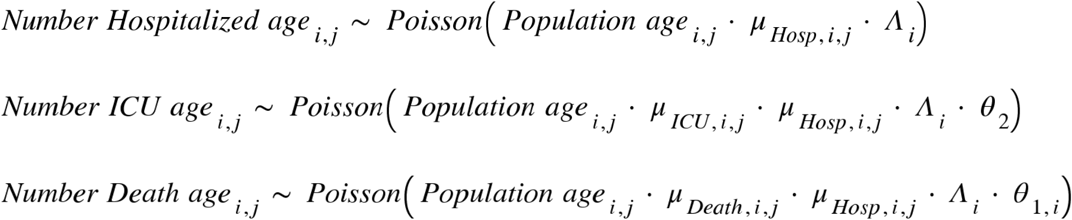

Where *μ*_*Hosp,i,j*_ is the probability of hospitalization for an individual within age group *i* of sex *j, μ*_*ICU,i,j*_ is the probability of entering ICU and *μ*_*Death,i,j*_ is the probability of death for hospitalized individuals within age group *i* of sex *j. θ*_2_ is the proportion of hospitalized individuals in the dataset that have experienced their ICU outcome and *θ*_1_,i is the proportion of individuals of age group *i* that have experienced their death outcome.

For the active surveillance portion of the model, we use a Poisson likelihood to capture the number of deaths on the Princess Diamond cruise ship. We assume that sufficient time has passed that all deaths that were going to occur by the passengers as a result of SARS-CoV-2 infection have now occurred. However, we conduct a sensitivity analysis where half of the remaining six patients that are still within ICU go on to die.

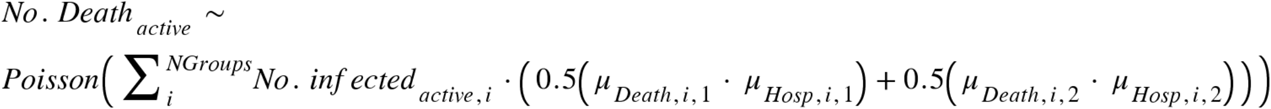

As this is a growing epidemic, many of the hospitalizations may yet end up being fatal. To adjust for this, we estimate the proportion of current hospitalizations where the outcome is known. *θ*_1,*i*_ represents the proportion of hospitalizations, where the death outcome is known for an individual of age *i*.

To estimate *θ*_1,*i*_, we calculate the proportion of hospitalized individuals who have already experienced their outcome (*18*).

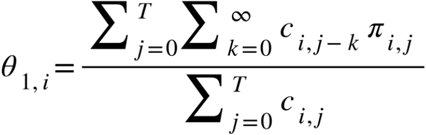

Where *C*_*i,j*_ is the number of cases at time *j* of age *i* and *π*_*i,j*_ is the proportion of all hospitalized cases in our dataset of age *i* that have a delay between hospitalization and death of *j* days.

We take a similar approach to estimate the proportion of hospitalized individuals who have experienced their ICU outcome (*θ*_2_).

We use RStan (*19*) to fit the *μ*_*Hosp,i,j*_, *μ*_*ICU,i,j*_, *μ*_*Death,i,j*_ and Λ parameters using logit transformed parameters. We run four chains of 10,000 iterations each and remove 50% for burn-in. We use 2.5% and 97.5% percentiles from the resulting posterior distributions for 95% credible intervals for the parameters. To calculate the overall probability of hospitalization following infection for the whole population we compute an average across the individual *μ*_*Hosp,i,j*_ estimates, weighted by the estimated number of people infected in each age-sex group. Similarly, to calculate the overall probability of death following hospitalisation, we compute an average across the individual *μ*_*Death,i,j*_ estimates, weighted by the estimated number of people hospitalised in each age-sex group.

### Transmission model fit to ICU data

We use a deterministic compartmental model stratified by age to describe the transmission of SARS-CoV-2 in the French population. Upon infection, susceptible individuals will enter a latent compartment (first exposed compartment *E*_1_), in which they will on average stay 4.0 days. During this period, they are not infectious. They will then move to a second exposed compartment *E*_2_ in which they will on average stay 1.0 day. Upon entry in the *E*_2_ compartment, infected individuals become infectious. They then move to the compartment *I* where they stay for an average duration of 3 days, where all individuals are infectious and a subset develop symptoms. This parametrization gives a mean incubation period of 5 days and allows for one day of pre-symptomatic transmissions, in the line with several estimates from Chinese data (*20, 21*). It is also in line with generation interval estimates obtained from analyses of infector-infectee pairs from mainland China (*20*).

A subset of infected individuals develop severe disease that results in ICU admission. The probability of ending up in ICU admission depends on age (*μ*_*Hosp,i,j*_ × *μ*_*ICU,i,j*_ for age i). Finally, we assume that patients will enter ICU on average 7 days after symptom onset, consistent with previous estimates (*22*).

The model is initiated with *I*_0_ cases in the *E*_1_ compartment on the 22nd January 2020 (*t*_0_).

### Contacts patterns in the French population prior to the lockdown

Age-specific daily contacts for the French population are obtained from the study COMES-F performed in 2012 (*7*). From this survey, we reconstruct the contact matrix describing mixing between age classes during a non-holiday period. To compute the matrix, we divide the population equally from 0 to 80 years old into 8 classes of ten years each. For the elderly, we consider one unique class that contains over-80 years old people. Daily contacts are computed by taking into account the variability associated with the weekend/weekdays seasonality. Data on contacts are retrieved and computed using the SocialmixR package (*23*).

### Computing the transmission rate prior to the lockdown

From the definition of the contact matrix, the parametrization of our transmission model and a given reproduction number *R*_0_, we can obtain the following expression for the transmission rate *β*(*24, 25*):

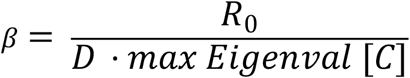

where *max Eigenval* [*C*] is the maximum eigenvalue of the contact matrix *C* and *D*is the mean infectious period.

### Trajectories of patients in ICU

We assume that the time spent in ICU is constant across age-groups and that it follows a Gamma distribution of shape 2 and of rate *g*^*out*^. This is modelled as two separate compartments for trajectories in ICU, from which individuals in ICU go out at rate *g*^*out*^. The mean time spent in ICU is thus equal to 2/*g* ^*out*^.

### Impact of the lockdown on transmission

In response to the growing epidemic, from March 17th, the French population was asked to remain confined to their homes and to avoid non-essential movement outside the household (*26*).

We adjusted our contact matrix to reflect the impact on the lockdown on the distribution of daily contacts between individuals after this date. We denote *C*, the contact matrix prior to the lockdown (*7*) and transmission rates prior to the lockdown are modelled as*βC*.

In order to model the impact of the lockdown on transmission, one potential approach is to predict how the standard contact matrix *C* is modified during the lockdown due to reductions in contacts in different settings. If we denote *CL* the predicted contact matrix during the lockdown period, the transmission rates for the lockdown period would then simply be *βCL*. However, a limitation of this approach is that given the unprecedented nature of the lockdown, it is hard to predict precisely what the new contact matrix *CL* may look like. Any slight error in the assumed reduction of the average number of contacts would have a strong impact on estimates of the reproduction number for the lockdown period.

To avoid such risk, we instead estimate a transmission parameter separately for the time period before (*β*) and during the lockdown (*β*_*lockdown*_). Comparison of these two parameters will determine the reduction in the reproduction number due to the lockdown. Since the reduction in average number of contacts will be captured by transmission parameters (*β, β*_*lockdown*_), we work with normalized contact matrices, i.e. contact matrices whose maximum eigenvalues are equal to 1. This allows us to define *β*as *R*_0_/*D* and *β*_*lockdown*_ as *R*_*lockdown*_/*D* and to compute transmission rates before and after lockdown as *βC* and *β*_*lockdown*_*CL*. We modify the contact matrix for the lockdown to capture the impact of the lockdown on the structure of the matrix. This normalization ensures that estimates of R after the lockdown are little impacted by the matrix we choose (see Figure 4).

The normalized contact matrices we consider for the lockdown matrices are:

- *CL*_1_ (baseline): the original contact matrix by removing all contacts in school settings and further assume a reduction of 80% in the contacts associated with the workplace and 90% in the ones outside work and home. This represents our baseline assumptions.
- *CL*_3_(Children Less Inf - children less infectious): same as *CL*_1_but where those aged <20 y.o. are 50% less infectious.
- *CL*_4_(CM No Change - contact matrix no change): the original (pre-lockdown) contact matrix (i.e. no change in the matrix).
- *CL*_5_(CM SDE - contact matrix social distancing elderly): same as *CL*_1_but with a further 60% reduction in all contacts of individuals aged over 70y.
- *CL*_5_(Constant AR - constant attack rates): all the coefficients of the contact matrix are equal to 1 (homogeneous mixing of the population).

We note that *CL*_3_and *CL*_4_are less likely to represent the true situation than *CL*_1_and*CL*_2_. *CL*_4_ would lead to only very few contacts for the elderly, and the model then predicts a substantial drop in the proportion of elderly individuals in ICU. Such reduction in ICU was not observed in France. Additionally, in the context of lockdowns that apply to the whole population, a recent study reports a fairly constant reduction in mean contacts across age classes from 18 years old and upwards before and after the lockdown (*27*)

### Statistical framework for the transmission model

We fit the transmission model using a Bayesian framework and jointly infer parameters. To do this we let *Adm*_*ICU*_^*pred*^ (*t*) and *ICU*^*pred*^ (*t*) denote respectively the number of admissions in ICU and the number of ICU beds on day *t*predicted by our model. We then let *Adm*_*ICU*_^*cor*^ (*t*) and *ICU*^*cor*^(*t*) respectively denote the corrected number of ICU admissions and the corrected number of ICU beds occupied on day *t*. The likelihood function until day *T*is:

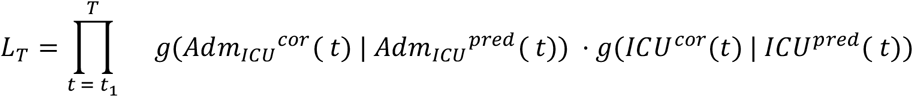

where *g*(. | *X*) is a negative binomial distribution of mean *X*and overdispersion parameter *X*^*δ*,^ *δ*being a parameter to be estimated. We calibrate the model on corrected SI-VIC data from the 15th of March (denoted *t*_1_).

The parameter space is explored by Markov Chain Monte Carlo sampling. We implement a Metropolis-Hastings (MH) algorithm with lognormal proposals for all the parameters and uniform priors. Chains are run for 10,000 iterations with 2,000 iterations of burn-in.

In early attempts to estimate model parameters, the initial number of cases at the start of the simulation *I*_0_ was highly correlated to the reproduction number. This is because slight variations in *I*_0_ or the reproduction number can lead to major changes in the trajectory of cumulative number of cases. We therefore re-parameterized the model to reduce this correlation by using a proxy for the number of incident cases at the time of the lockdown *I*_*lockdown*_.

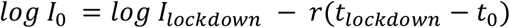

Where *I*_0_is the initial number of cases and r is the epidemic growth rate before the lockdown. We use the approach by Wallinga et al. (*28*) to relate the basic reproduction number to the epidemic growth rate *r*.

### Incorporation of uncertainty from the probability of entering ICu following uncertainty

We incorporate uncertainty from the probability of entering ICU following infection in our estimates of the number of new infections and the immunity in the population over time. To do this, we separately rerun the transmission model using the 2.5% and 97.5% quantiles from the posterior of *μ*_*Hosp,i,j*_ × *μ*_*ICU,i,j*_. The results of these estimates are included in Figures 3C, 3E, 4D, 4E and S4(A-M)3. Uncertainty in these parameters had little effect on our estimates of the number of required ICU beds and ICU admissions.

### Simulation study to assess model performance in estimating IFR and hospitalization risk

To assess the performance of the approach to estimate probabilities of infection, hospitalization, ICU entry, and death, we developed a simulation framework where the true parameters (*μ*_*Hosp,i,j*_, *μ*_*ICU,i,j*_,Λ, *μ*_*Death,i,j*_) were known.

For a period of 45 days we simulate a growing epidemic, seeded by a single infection, where the number of cases grows exponentially each day with an exponential growth rate of 0.3. We assume a population with the same age structure as France and assume no difference in risk of infection by age or sex. For each of the infections in the simulation we assign:

- The age group, *i*, drawn according to the age distribution of France
- Whether or not the individual was hospitalized, using a random draw from a Bernoulli distribution with parameter *μ*_*Hosp,i,j*_
- If the individual was hospitalized, whether or not the individual entered ICU, using a random draw from a Bernoulli distribution with parameter *μ*_*ICU,i,j*_
- If the individual was hospitalized, whether or not the individual died, using a random draw from a Bernoulli distribution with parameter *μ*_*Death,i,j*_
- If the individual was hospitalized, the day of hospitalization using an exponential distribution with a mean of 11 days.
- If the individual entered ICU, the delay from hospitalization to ICU using a random draw from an exponential distribution with a mean of 2 days.
- If the individual died, the delay from hospitalization to death using a random draw from an exponential distribution with a mean of 15 days.

We then compute the total counts of hospitalizations, ICU and deaths by age over the first 45 days of the simulation (Figures S7).

To simulate active surveillance, we select a random subset of 1000 individuals that were infected and record the outcome (death or not) and age for all of them (irrespective of delays to death).

We use the simulated data to estimate the proportion of cases with outcome observed (*θ*) and the model parameters using our probabilistic framework.

## Data Availability

All data than can be made available will be put on a GitHub link

## Supplementary Information

### Supplementary tables

**Table S1:**
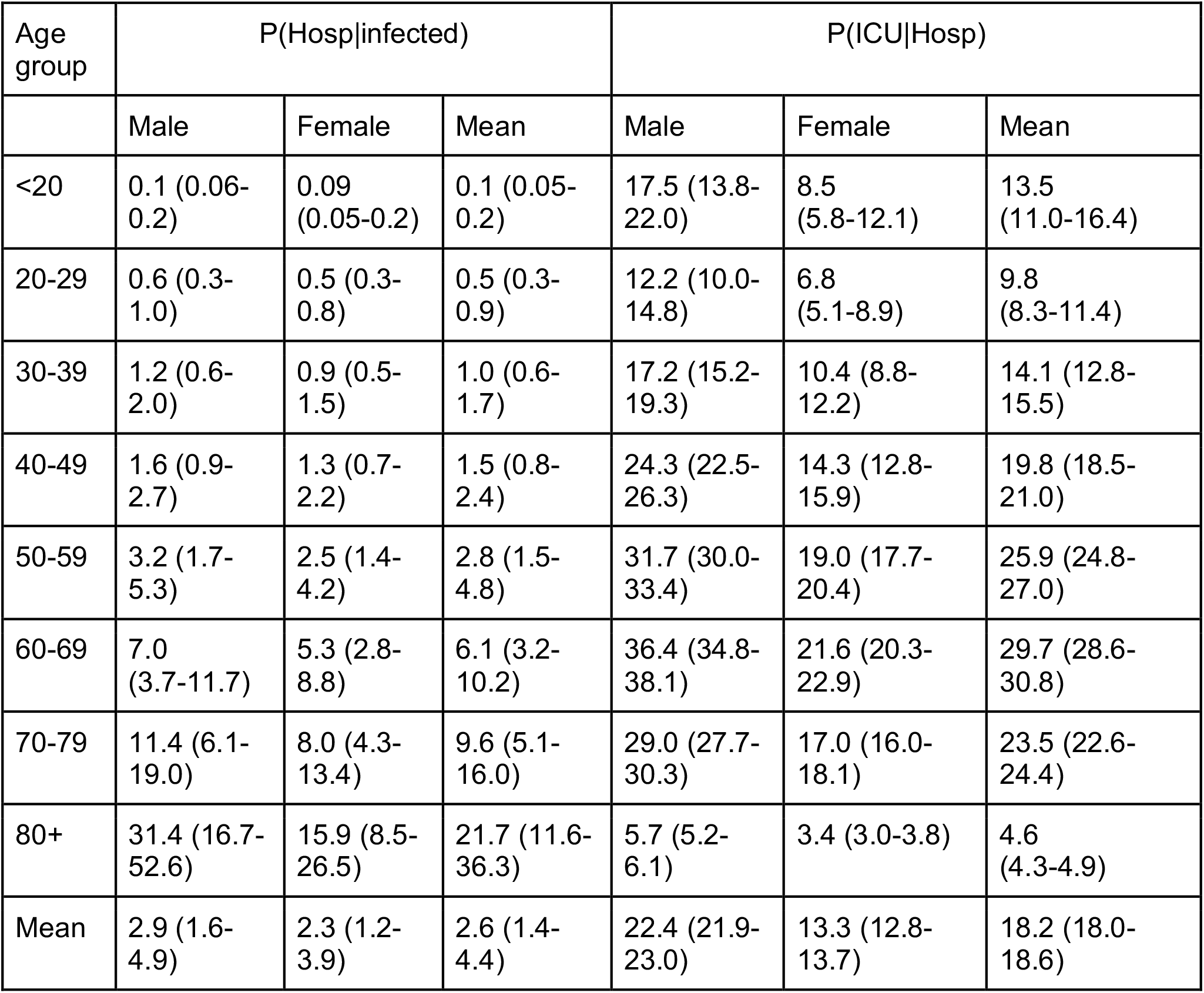
Probability of hospitalization and ICU by age and sex.

**Table S2:** Probability of death by age and sex.

| Age group | P(Death\|Hosp) |  |  | Infection fatality proportion |  |  |
| --- | --- | --- | --- | --- | --- | --- |
|  | Male | Female | Mean | Male | Female | Mean |
| <20 | 1.2 (0.4-2.8) | <0.001 | 0.6 (0.2-1.5) | 0.001 (<0.001-0.004) | <0.001 | 0.001 (0.000-0.002) |
| 20-29 | 1.3 (0.6-2.4) | 1.4 (0.6-2.7) | 1.4 (0.8-2.2) | 0.007 (0.003-0.02) | 0.007 (0.002-0.02) | 0.007 (0.003-0.01) |
| 30-39 | 2.5 (1.8-3.4) | 1.6 (0.9-1.4) | 2.1 (1.6-2.7) | 0.03 (0.01-0.05) | 0.01 (0.006-0.03) | 0.02 (0.01-0.04) |
| 40-49 | 3.9 (3.1-4.7) | 3.2 (2.5-4.1) | 3.6 (3.0-4.2) | 0.06 (0.03-0.1) | 0.04 (0.02-0.07) | 0.05 (0.03-0.09) |
| 50-59 | 7.5 (6.6-8.3) | 6.4 (5.6-7.2) | 7.0 (6.4-7.6) | 0.2 (0.1-0.4) | 0.2 (0.08-0.3) | 0.2 (0.1-0.3) |
| 60-69 | 14.2 (16.2-15.3) | 12.0 (11.0-13.1) | 13.2 (12.5-13.9) | 1.0 (0.5-1.7) | 0.6 (0.3-1.1) | 0.8 (0.4-1.4) |
| 70-79 | 25.3 (24.1-26.6) | 20.7 (19.5-22.0) | 23.2 (22.3-24.1) | 2.9 (1.5-4.8) | 1.7 (0.9-2.8) | 2.2 (1.2-3.7) |
| 80+ | 42.0 (40.7-43.4) | 34.0 (32.7-35.4) | 38.4 (37.4-39.3) | 13.2 (7.0-22.1) | 5.4 (2.9-9.1) | 8.3 (4.4-13.9) |
| Mean | 21.8 (21.3-22.3) | 17.8 (17.3-18.4) | 20.0 (19.6-20.4) | 0.6 (0.3-1.1) | 0.4 (0.2-0.7) | 0.5 (0.3-0.9) |

**Table S3:** Estimated delays from hospitalization to death.

| Age group | Parameters |  |  |  | Overall Mean (days) |
| --- | --- | --- | --- | --- | --- |
|  | P(short delay) | Exponential<br>(for short delay) | Lognormal<br>(for longer delays) |  |  |
|  |  | Mean (days) | Mean (days) | Median (days) |  |
| <70 | 0.11 | 0.67 | 21.2 | 12.4 | 14.0 |
| 70-80 | 0.13 | 0.67 | 12.6 | 8.5 | 10.3 |
| 80+ | 0.18 | 0.67 | 10.5 | 7.5 | 8.6 |
| Mean | 0.15 | 0.67 | 13.2 | 8.6 | 10.1 |

**Table S4:** Parameter estimates from the national model.

| Parameter | Estimate with 95% credible interval |
| --- | --- |
| Basic reproduction number $R_0$ | 3.31 [3.18 - 3.43] |
| Reproduction number after lockdown $R_{lockdown}$ | 0.52 [0.5 - 0.55] |
| Overdispersion parameter $\delta$ | 0.77 [0.7 - 0.83] |
| Initial number of cases $I_0$ | 15.83 [9.87 - 26.34] |
| Mean time spent in ICU $2/g^{out}$ | 17.15 [16.24 - 18.3] |

**Table S5:**
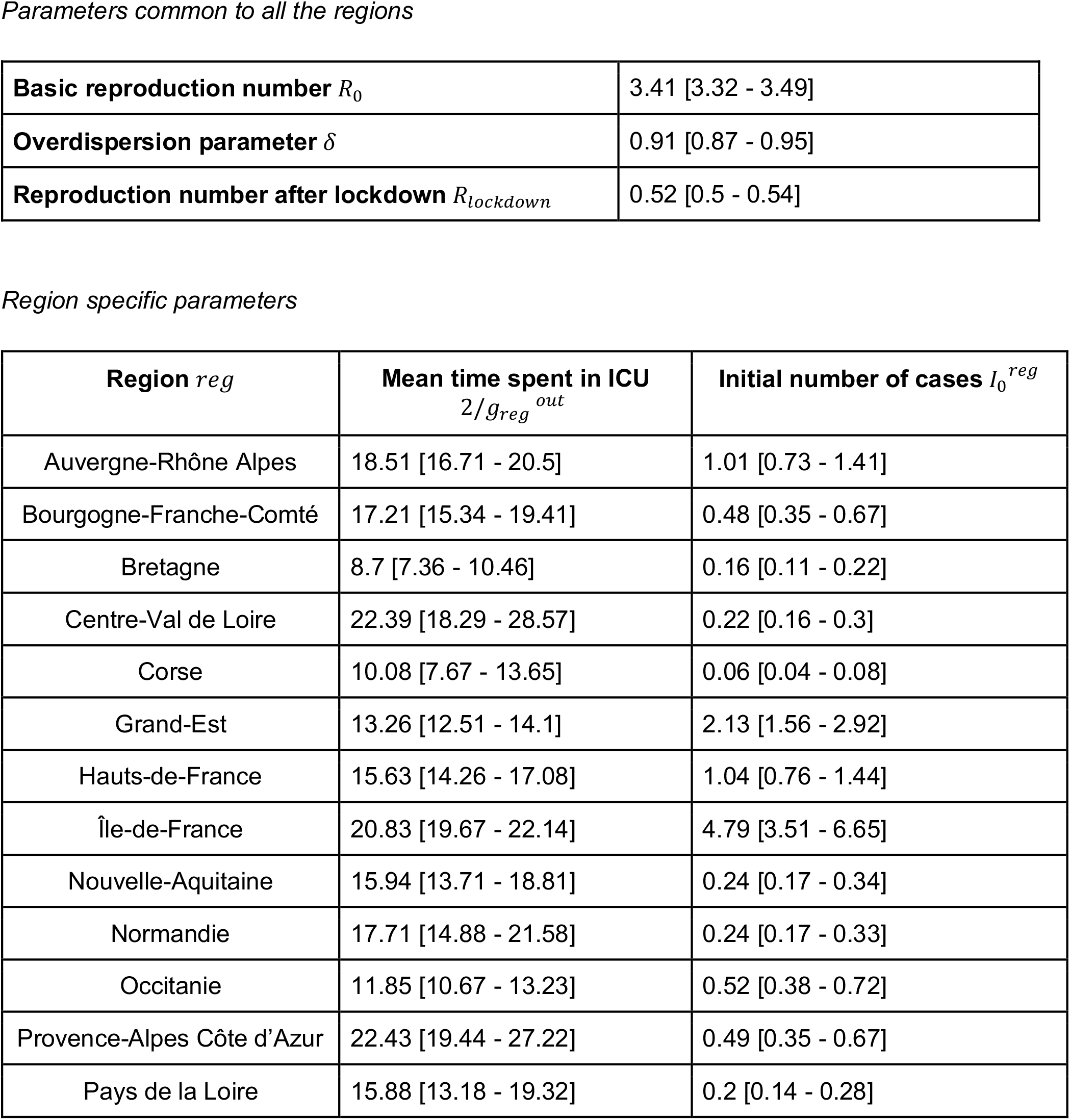
Parameter estimates from the regional model.

**Table S6:** Proportion infected by region by the 11th May.

| **Region** | **Proportion infected (%) (with 95% uncertainty range stemming from the uncertainty in the probability of entering ICU following infection)** |
| --- | --- |
| Auvergne-Rhône Alpes | 4.4 [2.7 - 8.3] |
| Bourgogne-Franche-Comté | 5.7 [3.5 - 10.6] |
| Bretagne | 1.8 [1.1 - 3.3] |
| Centre-Val de Loire | 3.1 [1.9 - 5.8] |
| Corse | 5.4 [3.3 - 10.2] |
| Grand-Est | 11.8 [7.4 - 20.5] |
| Hauts-de-France | 6.1 [3.7 - 11.3] |
| Île-de-France | 12.3 [7.9 - 21.3] |
| Nouvelle-Aquitaine | 1.4 [0.9 - 2.8] |
| Normandie | 2.6 [1.5 - 4.9] |
| Occitanie | 3.1 [1.9 - 5.9] |
| Provence-Alpes Côte d'Azur | 3.4 [2.1 - 6.4] |
| Pays de la Loire | 1.9 [1.2 - 3.8] |

### Supplementary figures

**Figure S1:**
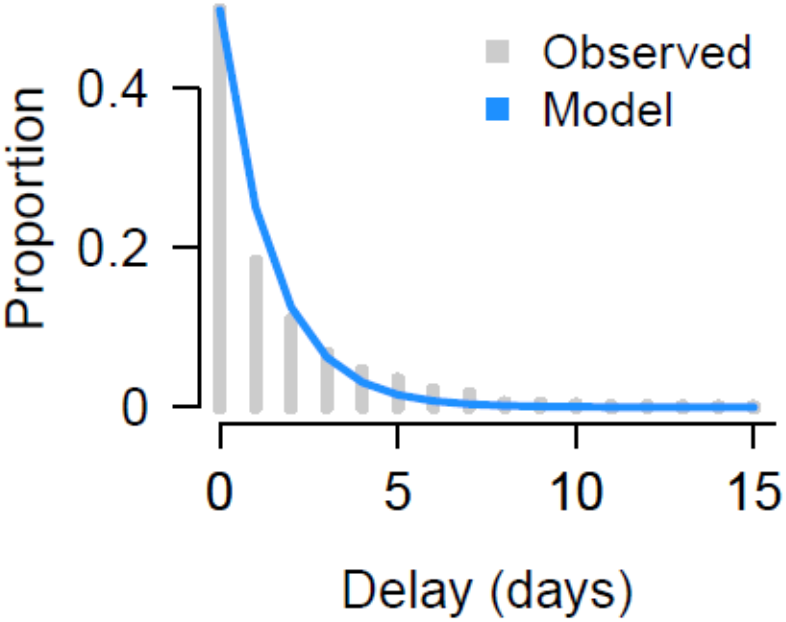
Fit of delay from hospitalization to ICU admission. and observed fit of exponential model use times from hospitalization to ICU entry across all ages, taking account for the exponentially growing nature of the epidemic.

**Figure S2:**
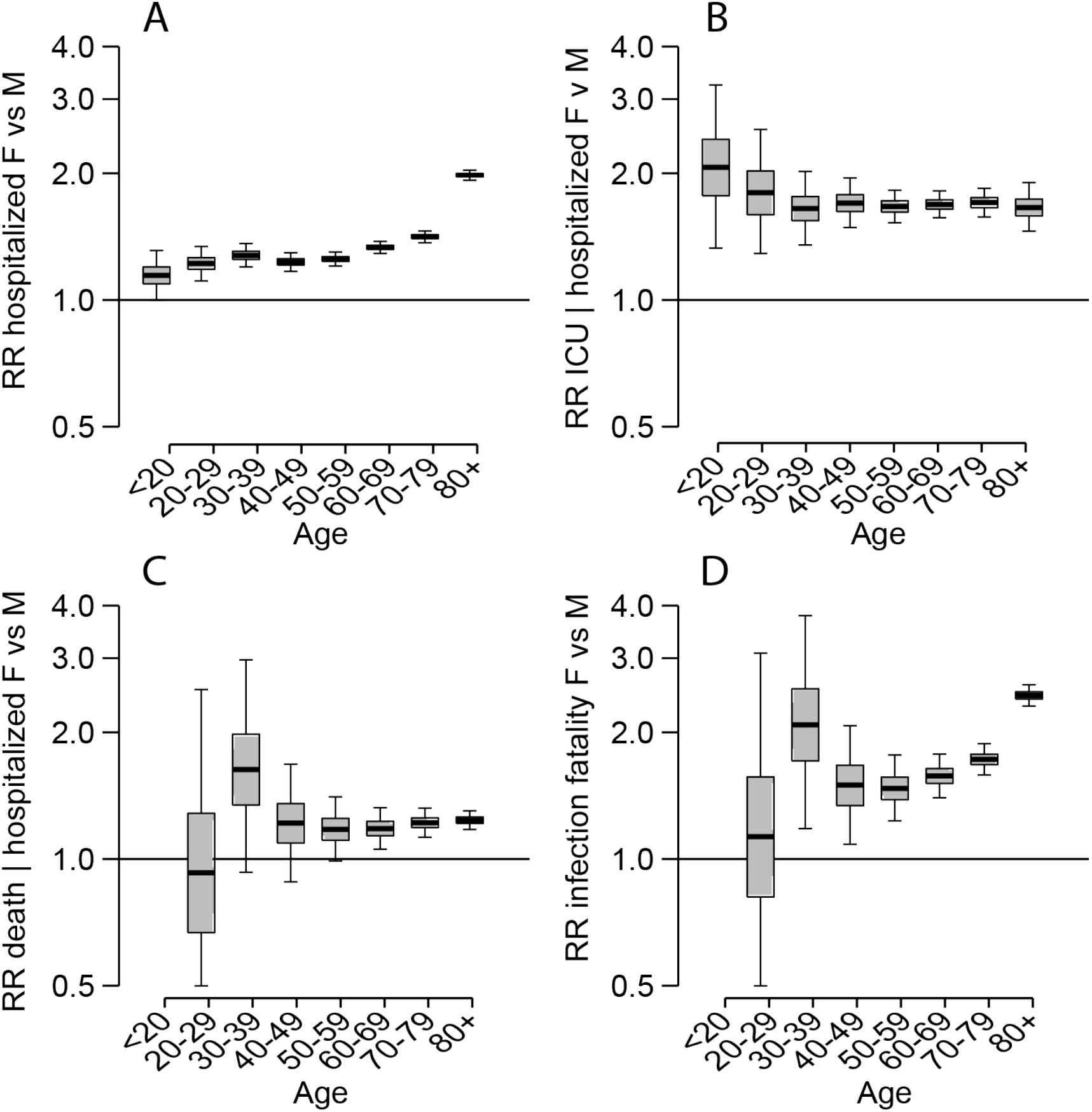
Relative differences by sex. **(A)** Relative risk of hospitalization. **(B)** Relative risk of ICU entry given hospitalization,**(C)** Relative risk of death among those hospitalized. **(D)** Relative risk of death among all those infected.

**Figure S3:**
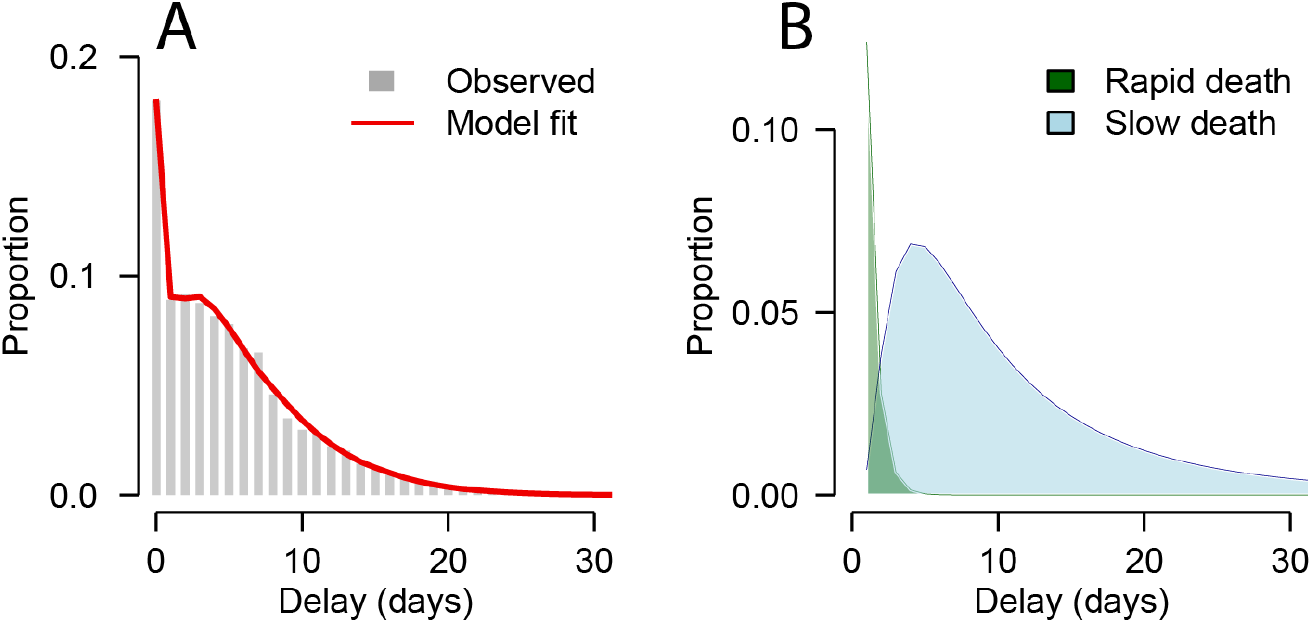
Fit of delays from hospitalization to death. **(A)** Observed and fitted distribution of delays between hospital admission and death. **(B)** Model estimates of distribution of rapid decline and slow decline. Models fitted to take into account that in a growing epidemic, observed deaths will be biased towards ones that die quickly.

**Figure S4:**
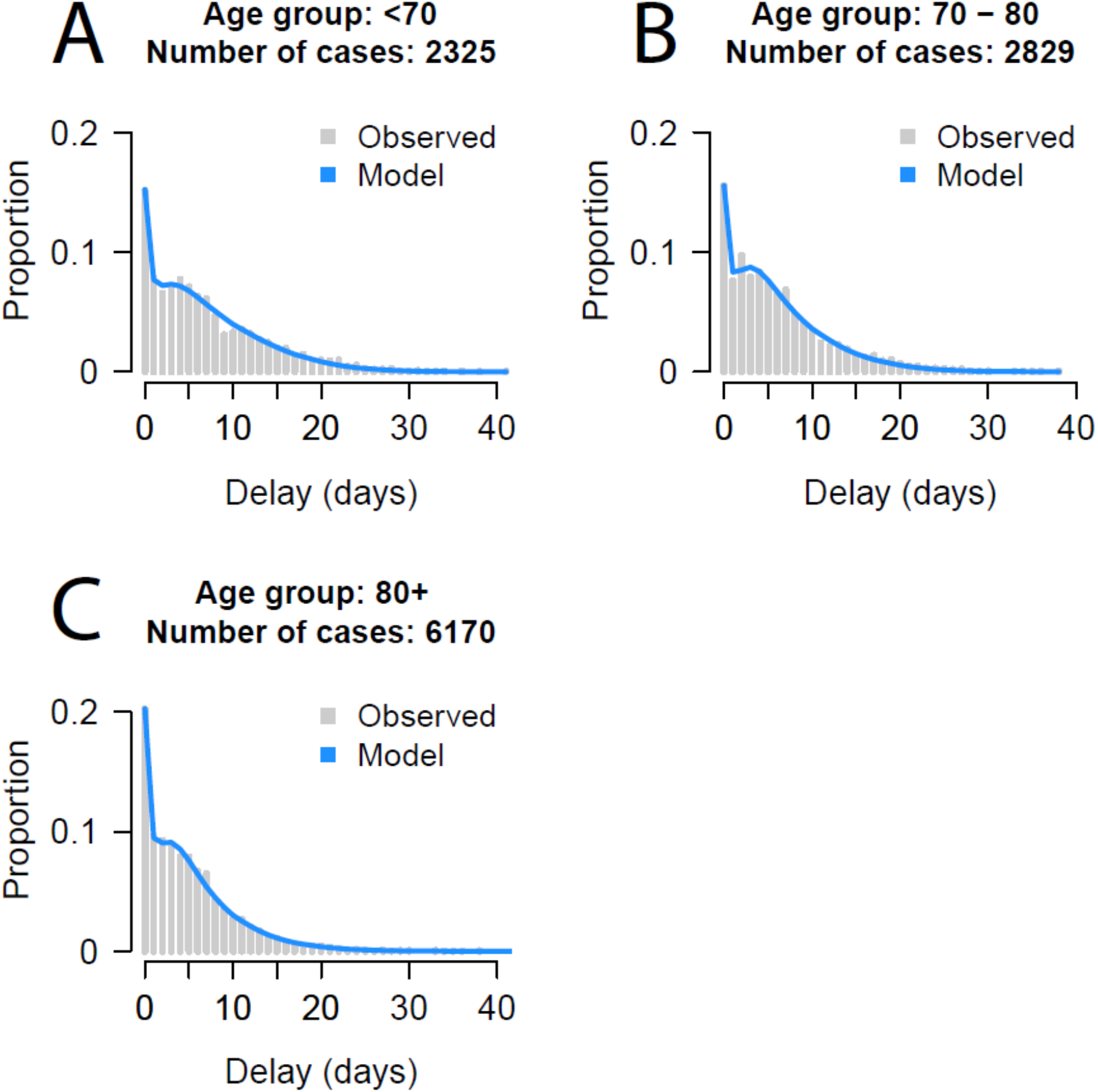
Fit of delays from hospitalization to death by age. Fit of mixture models to time from hospitalization to death for different age groups. The models are mixture models that have both an exponential decay for those that die quickly and a log-normal component for those that die after longer delays.

**Figure S5:**
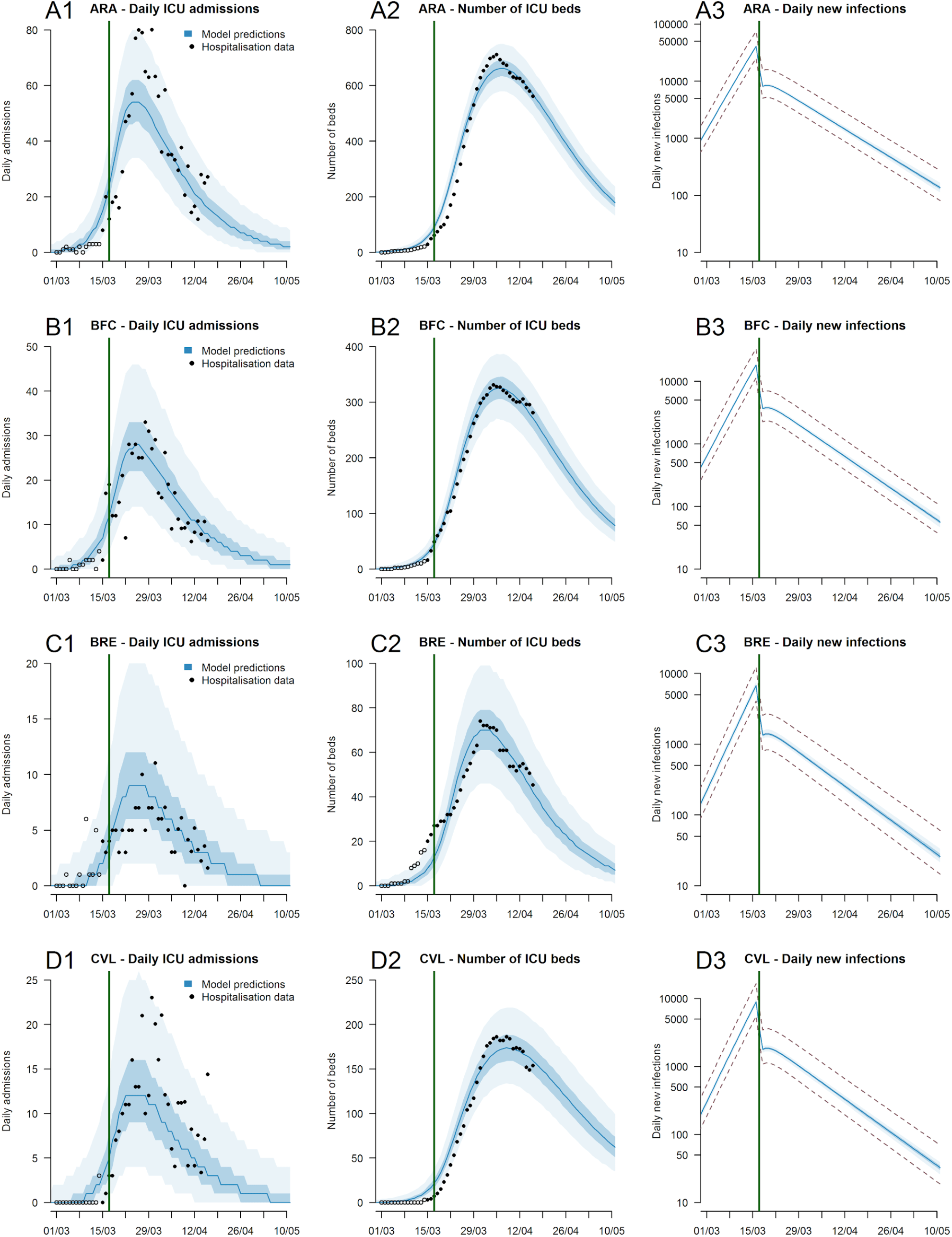
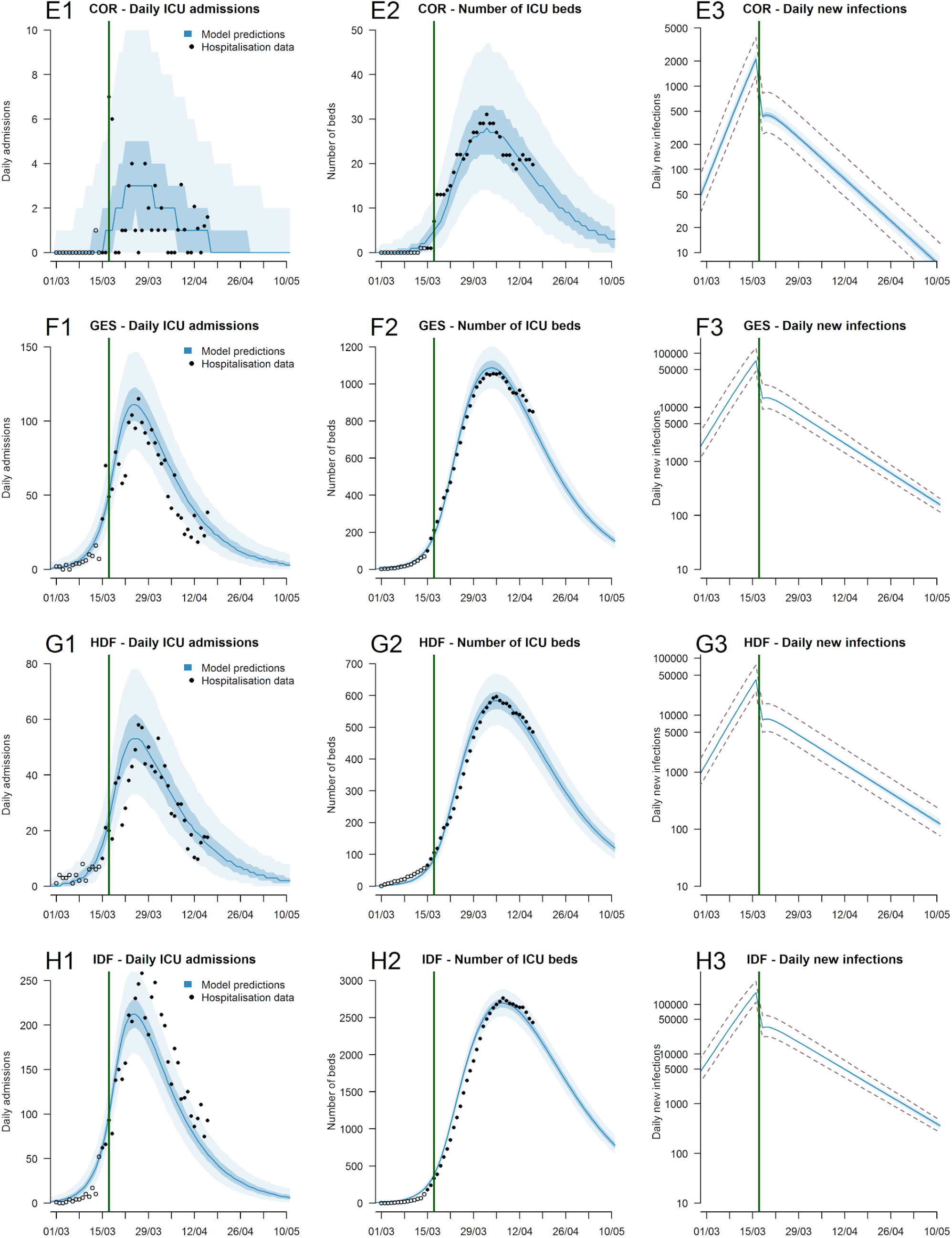
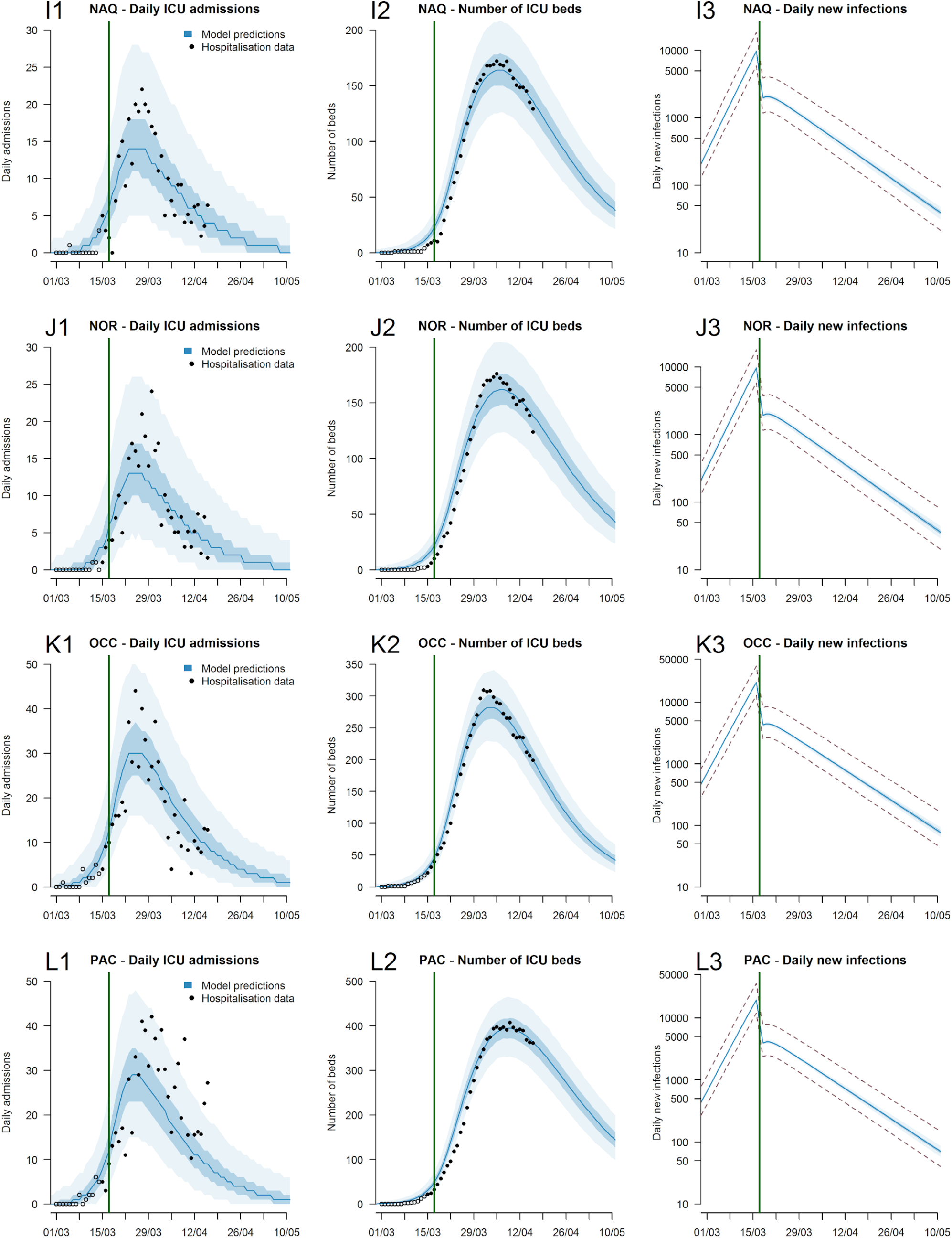
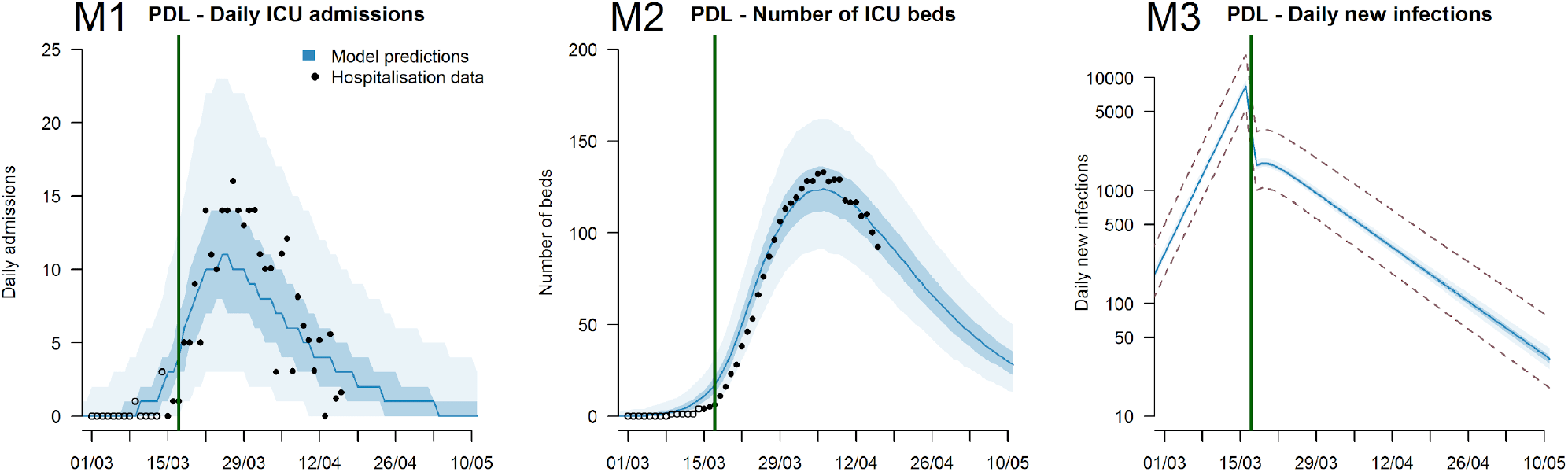
Trajectories predicted by the regional model. Predictions per French region (A) Auvergne-Rhône-Alpes; (B) Bourgogne-Franche-Comté : (C) Bretagne : (D) Centre-Val de Loire; (E) Corse; (F) Grand-Est; (G) Hauts-de-France; (H) Ïle-de-France; (I) Nouvelle-Aquitaine; (J) Normandie; (K) Occitanie; (L) Provence-Alpes Côte d’Azur; (M) Pays-de la Loire. (1) : Daily ICU admissions. (2) Number of ICU beds (3) Daily number of infections (logarithmic scale). The green line indicates the time intervention measures were put in place that limited movement in the country. The dotted lines in panels 3 represent the 95% uncertainty range stemming from the uncertainty in the probability of entering ICU following infection.

**Figure S6:**
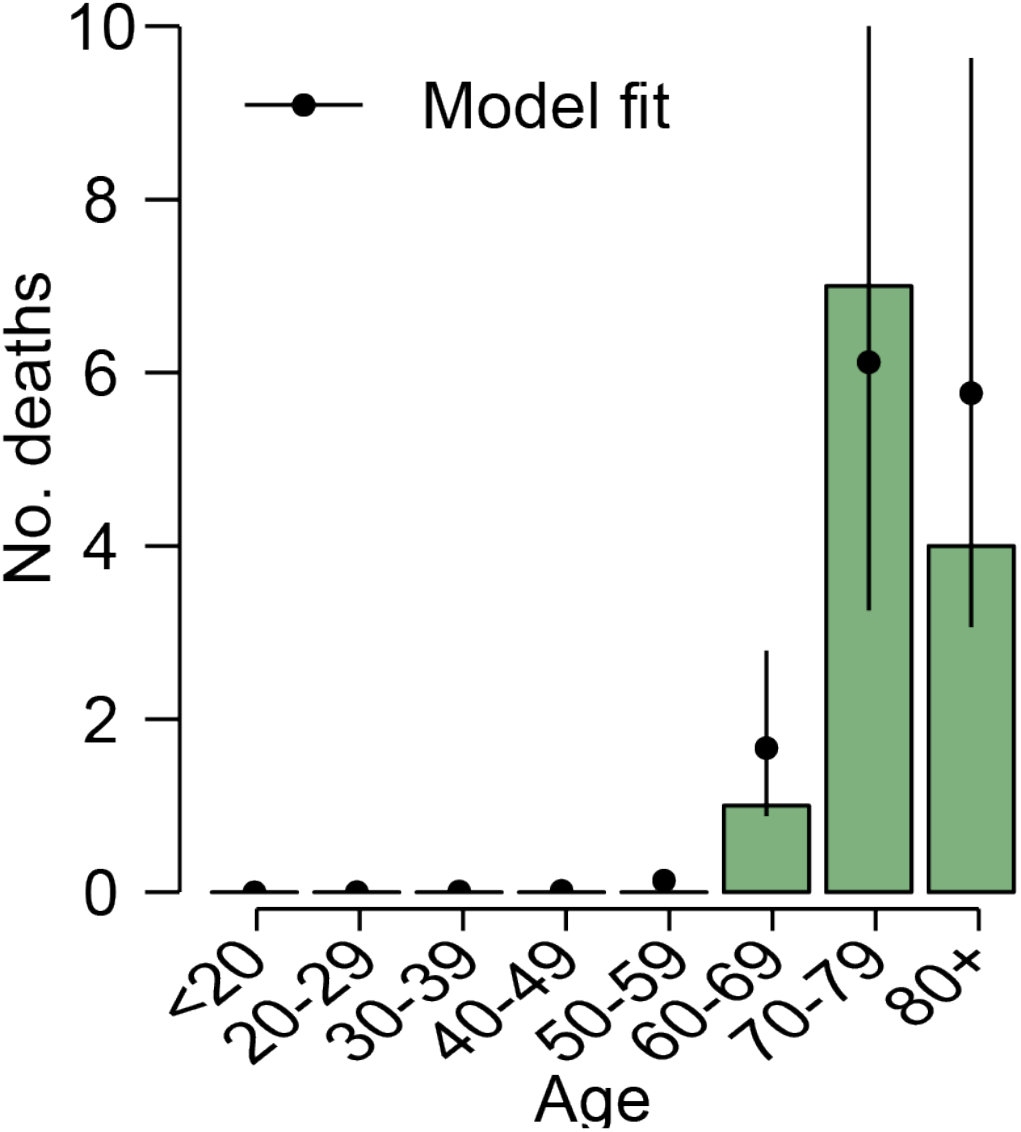
Princess Diamond fit. The observed (green bars) and fitted (black line) number of deaths from passengers on board the Princess Diamond who were infected with SARS-CoV-2. Note there is one fatal case where no age was reported and is therefore excluded from the plot (their death is included in the model estimation).

**Figure S7:**
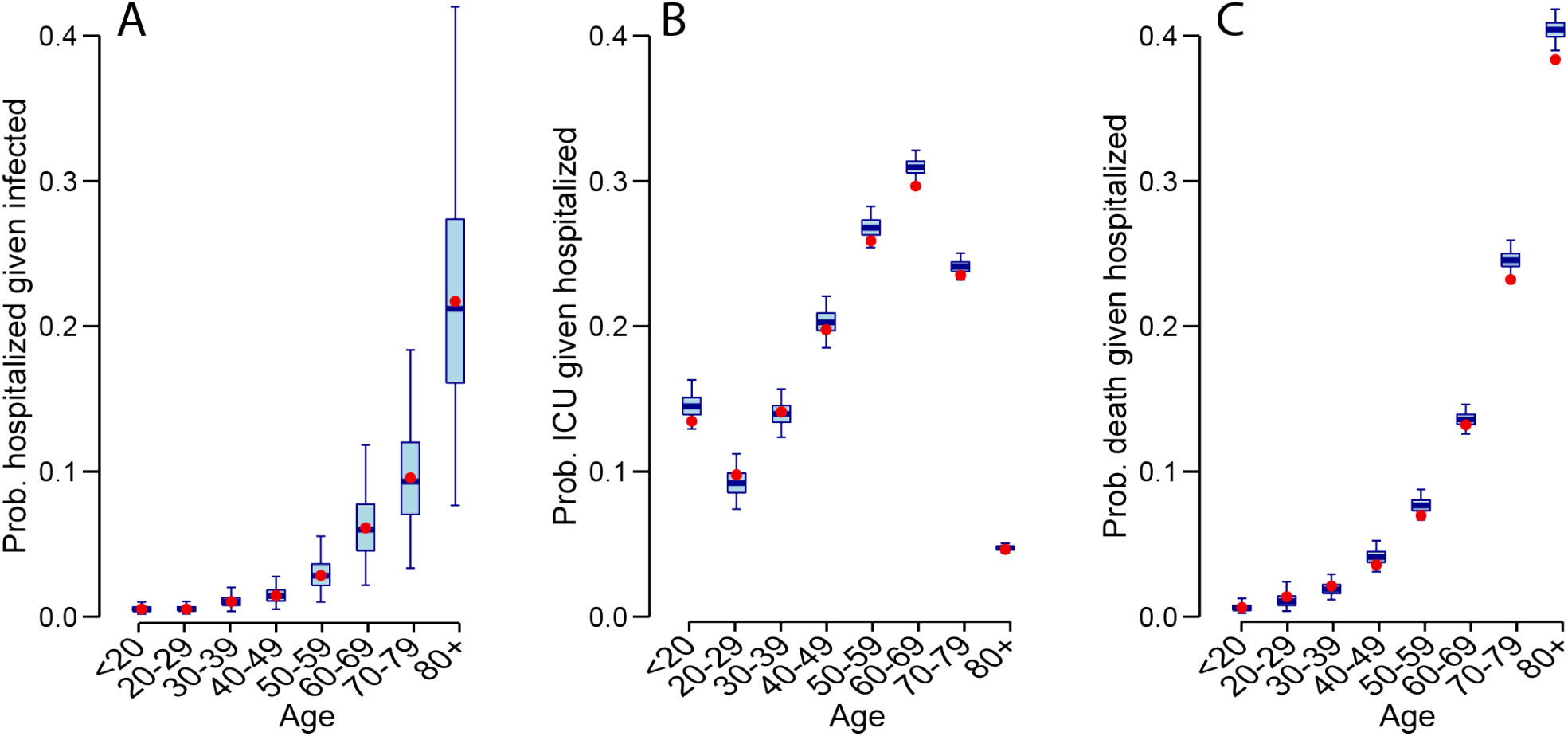
Simulation results. Simulation results where epidemics are simulated with known probabilities of infection, hospitalization, ICU and death. We then use our model framework to re-estimate the parameters. **(A)** Estimated (blue) and true (red) probability of hospitalization by age. **(B)** Estimated (blue) and true (red) probability of ICU admission by age. **(C)** Estimated (blue) and true (red) probability of death by age among those hospitalized.

**Figure S8:**
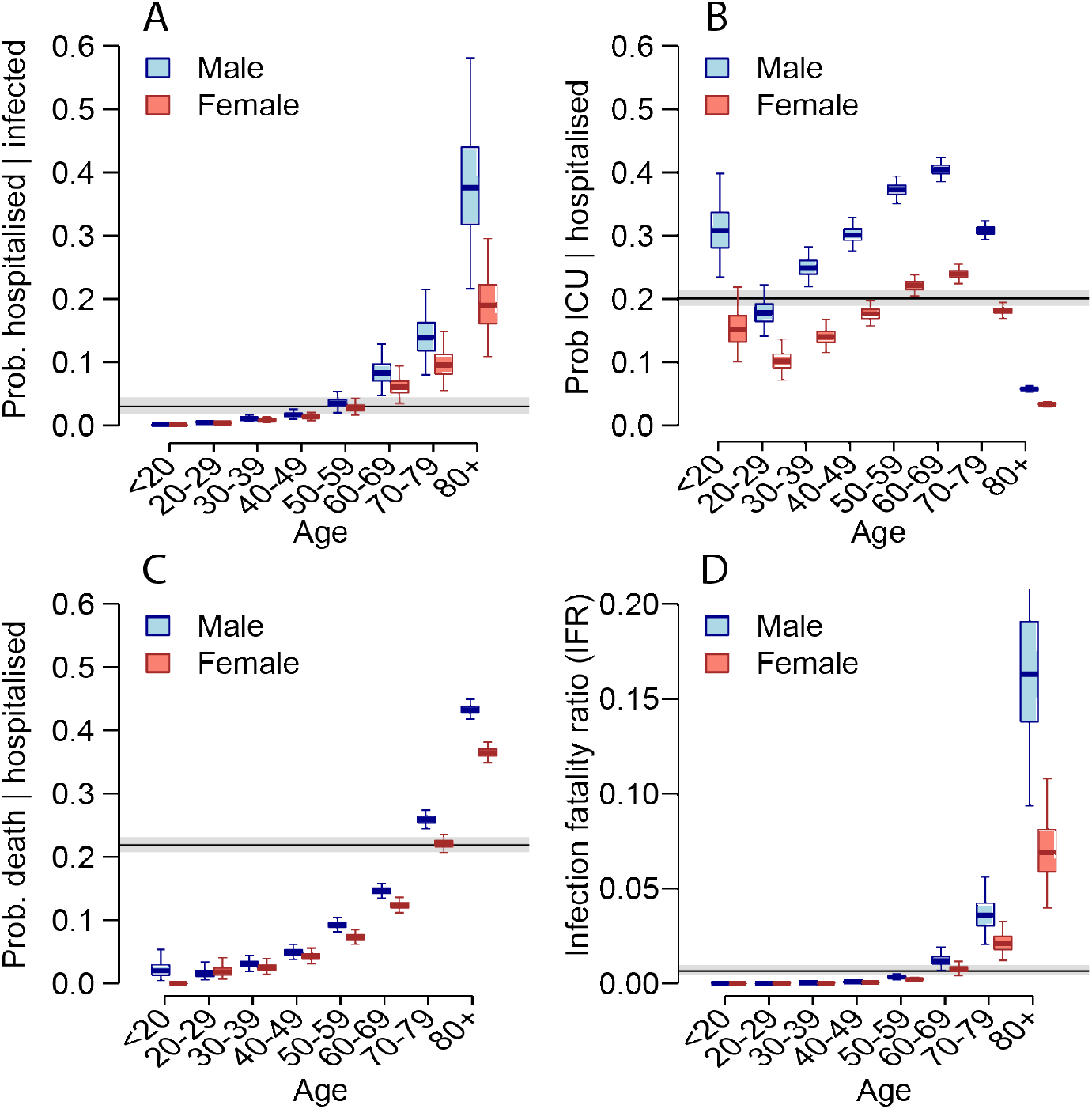
Sensitivity analysis with increased deaths on Princess Diamond. Six infected individuals remain in ICU. This sensitivity analysis assumes that half of these will go on to die. **(A)** Probability of hospitalization among those infected as a function of age and sex. **(B)** Probability of ICU admission among those hospitalized as a function of age and sex. **(C)** probability of death among those hospitalized as a function of age and sex. **(D)** Probability of death among those infected as a function of age and sex. For each panel, the black line and grey shaded region represents the overall mean across all ages.

**Figure S9:**
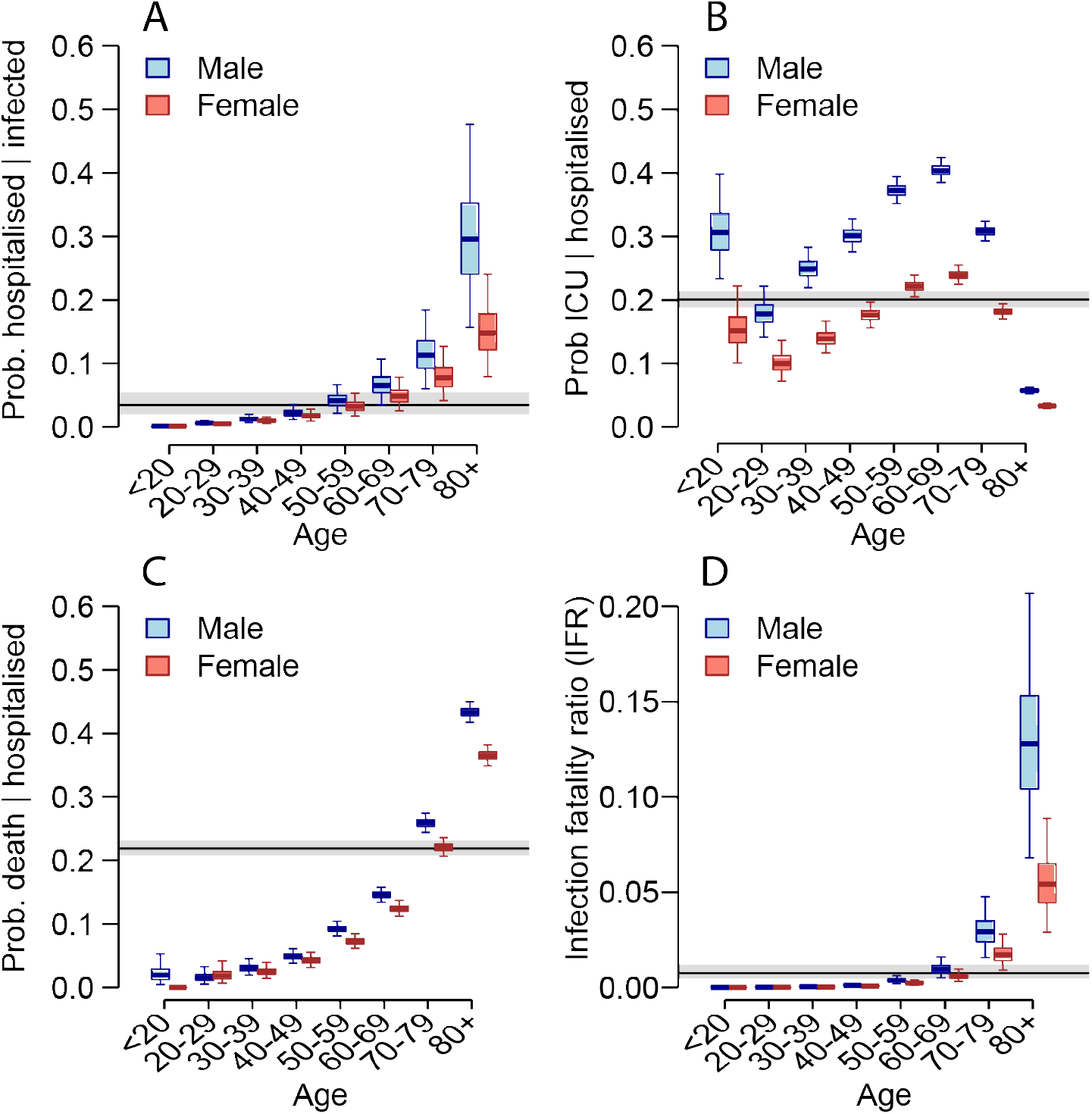
Sensitivity analysis with equal attack rates across age groups. **(A)** Probability of hospitalization among those infected as a function of age and sex. **(B)** Probability of ICU admission among those hospitalized as a function of age and sex. **(C)** Probability of death among those hospitalized as a function of age and sex. **(D)** Probability of death among those infected as a function of age and sex. For each panel, the black line and grey shaded region represents the overall mean across all ages.

**Figure S10:**
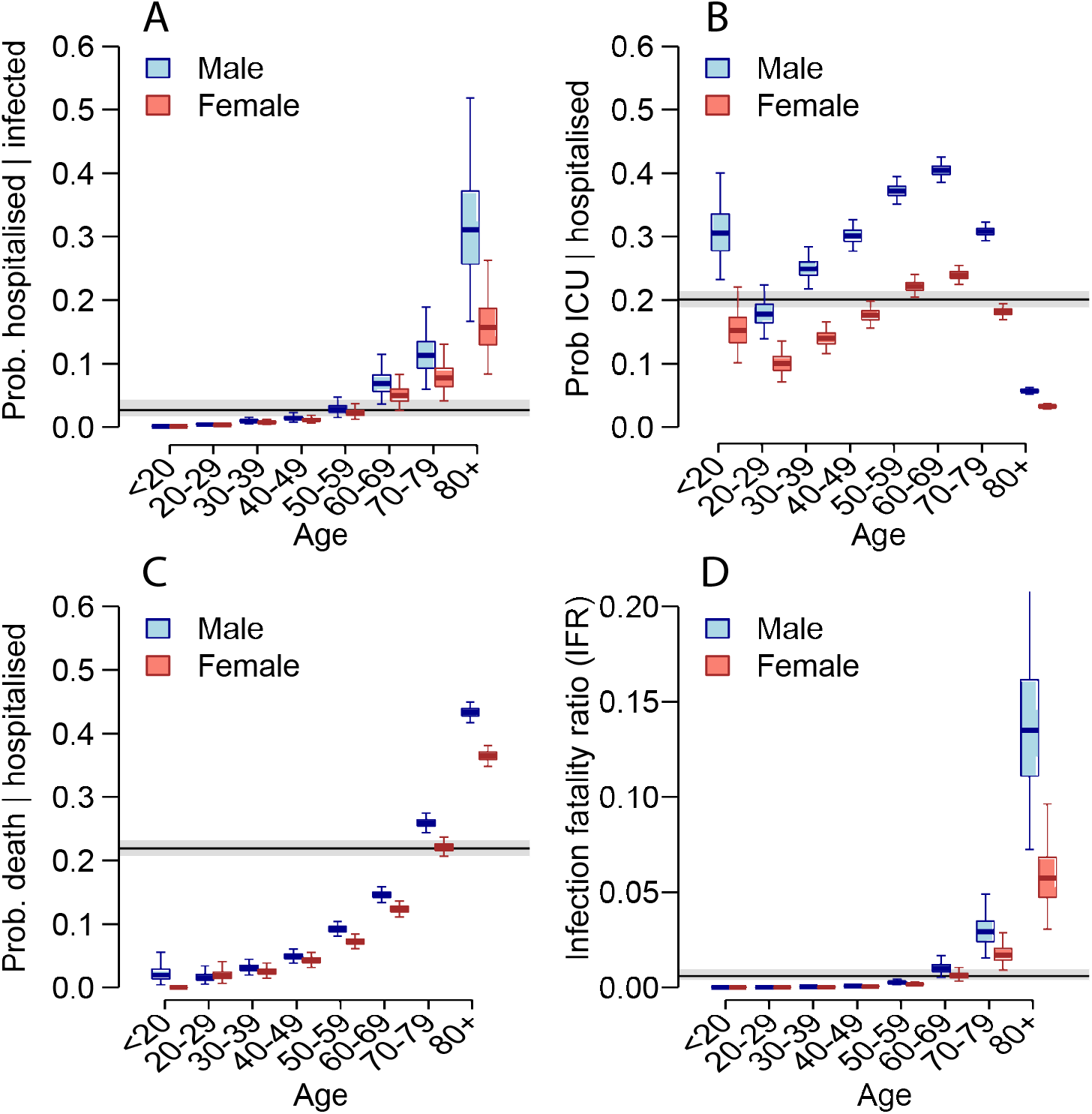
Sensitivity analysis with reduced transmission in <20y. Sensitivity analysis where we explore impact of increased levels of asymptomatic infection may result in reduced transmission among those <20y. **(A)** Probability of hospitalization among those infected as a function of age and sex. **(B)** Probability of ICU admission among those hospitalized as a function of age and sex. **(C)** Probability of death among those hospitalized as a function of age and sex. **(D)** Probability of death among those infected as a function of age and sex. For each panel, the black line and grey shaded region represents the overall mean across all ages.

**Figure S11:**
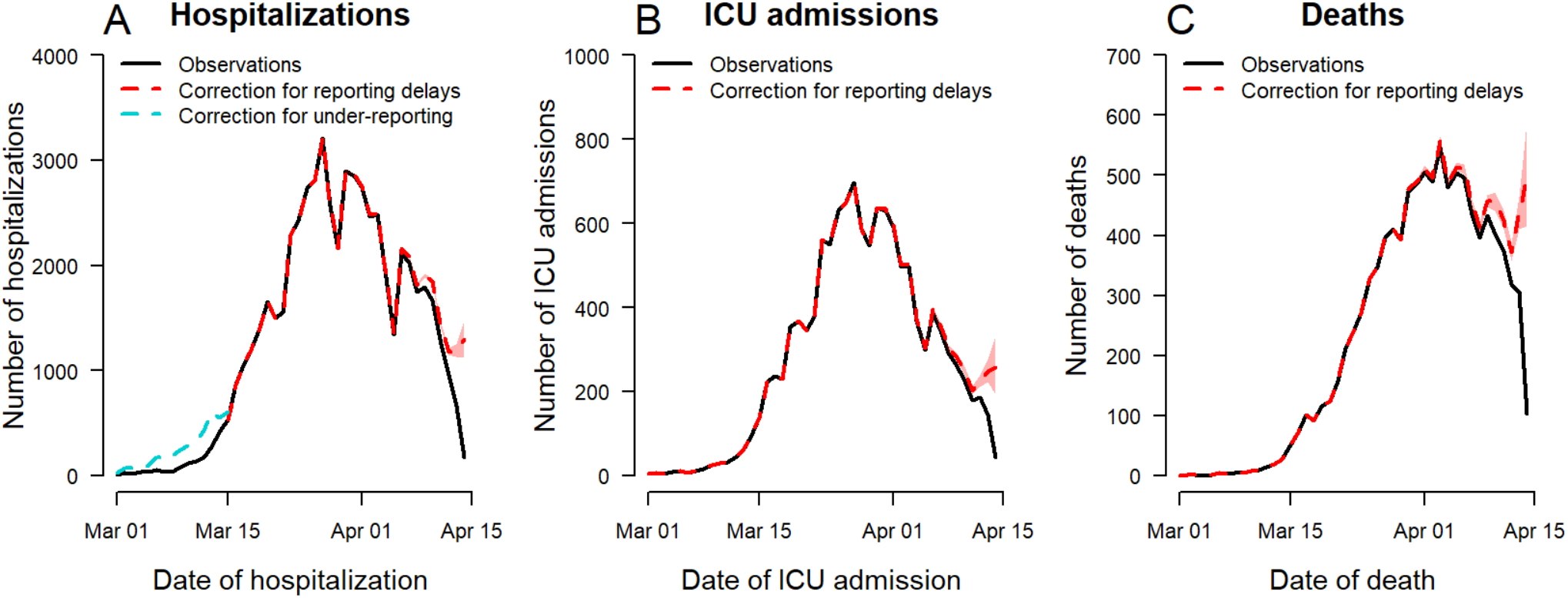
Time-series of hospitalizations, ICU admissions and deaths corrected for reporting delays. Times-series of hospitalizations (A), ICU admissions (B) and deaths (C) from SI-VIC data, corrected for reporting delays and under-reporting. The SI-VIC system became operational on the 13th of March. Deaths and ICUs were retrospectively added, however some hospitalizations that occurred prior to or around this date were likely missed. To account for missing hospitalizations prior to the 15th of March, we used another reporting system (OSCOUR®) that was already established (blue line in (A)). We also account for delays in reporting to estimate the number of hospitalizations, ICU admissions and deaths at the right end of the curves (red line in each panel). See methods section on how these were calculated.

## References

1. M. U. G. Kraemer, C.-H. Yang, B. Gutierrez, C.-H. Wu, B. Klein, D. M. Pigott, Open COVID-19 Data Working Group, L. du Plessis, N. R. Faria, R. Li, W. P. Hanage, J. S. Brownstein, M. Layan, A. Vespignani, H. Tian, C. Dye, O. G. Pybus, S. V. Scarpino, The effect of human mobility and control measures on the COVID-19 epidemic in China. Science (2020), doi:10.1126/science.abb4218.

2. J. Lourenço, R. Paton, M. Ghafari, M. Kraemer, C. Thompson, P. Simmonds, P. Klenerman, S. Gupta, Fundamental principles of epidemic spread highlight the immediate need for large-scale serological surveys to assess the stage of the SARS-CoV-2 epidemic. medRxiv (2020) (available at https://www.medrxiv.org/content/10.1101/2020.03.24.20042291v1.abstract).

3. L. Bao, W. Deng, H. Gao, C. Xiao, J. Liu, J. Xue, Q. Lv, J. Liu, P. Yu, Y. Xu, F. Qi, Y. Qu, F. Li, Z. Xiang, H. Yu, S. Gong, M. Liu, G. Wang, S. Wang, Z. Song, W. Zhao, Y. Han, L. Zhao, X. Liu, Q. Wei, C. Qin, Reinfection could not occur in SARS-CoV-2 infected rhesus macaques,, doi:10.1101/2020.03.13.990226.

4. J. Lessler, H. Salje, M. D. Van Kerkhove, N. M. Ferguson, S. Cauchemez, I. Rodriquez-Barraquer, R. Hakeem, T. Jombart, R. Aguas, A. Al-Barrak, D. A. T. Cummings, MERS-CoV Scenario and Modeling Working Group, Estimating the Severity and Subclinical Burden of Middle East Respiratory Syndrome Coronavirus Infection in the Kingdom of Saudi Arabia. Am. J. Epidemiol. 183, 657–663 (2016).

5. R. Verity, L. C. Okell, I. Dorigatti, P. Winskill, C. Whittaker, N. Imai, G. Cuomo-Dannenburg, H. Thompson, P. G. T. Walker, H. Fu, A. Dighe, J. T. Griffin, M. Baguelin, S. Bhatia, A. Boonyasiri, Cori, Z. Cucunubá, R. FitzJohn, K. Gaythorpe, W. Green, A. Hamlet, W. Hinsley, D. Laydon, G. Nedjati-Gilani, S. Riley, S. van Elsland, E. Volz, H. Wang, Y. Wang, X. Xi, C. A. Donnelly, A. C. Ghani, N. M. Ferguson, Estimates of the severity of coronavirus disease 2019: a model-based analysis. Lancet Infect. Dis. (2020), doi:10.1016/S1473-3099(20)30243-7.

6. P. G. T. Walker, C. Whittaker, O. Watson, M. Baguelin, K. E. C. Ainslie, S. Bhatia, S. Bhatt, A. Boonyasiri, O. Boyd, L. Cattarino, Others, The global impact of covid-19 and strategies for mitigation and suppression. On behalf of the imperial college covid-19 response team, Imperial College of London (2020).

7. G. Béraud, S. Kazmercziak, P. Beutels, D. Levy-Bruhl, X. Lenne, N. Mielcarek, Y. Yazdanpanah, P.-Y. Boëlle, N. Hens, B. Dervaux, The French Connection: The First Large Population-Based Contact Survey in France Relevant for the Spread of Infectious Diseases. PLoS One. 10, e0133203 (2015).

8. T. W. Russell, J. Hellewell, C. I. Jarvis, K. van Zandvoort, S. Abbott, R. Ratnayake, Cmmid Covid-Working Group, S. Flasche, R. M. Eggo, W. J. Edmunds, A. J. Kucharski, Estimating the infection and case fatality ratio for coronavirus disease (COVID-19) using age-adjusted data from the outbreak on the Diamond Princess cruise ship, February 2020. Euro Surveill. 25 (2020), doi:10.2807/1560-7917.ES.2020.25.12.2000256.

9. K. Mizumoto, K. Kagaya, G. Chowell, Early epidemiological assessment of the transmission potential and virulence of coronavirus disease 2019 (COVID-19) in Wuhan City: China, January-February, 2020. medRxiv (2020) (available at https://www.medrxiv.org/content/10.1101/2020.02.12.20022434v2.abstract).

10. L. Peeples, News Feature: Avoiding pitfalls in the pursuit of a COVID-19 vaccine. Proc. Natl. Acad. Sci. U. S. A. (2020), doi:10.1073/pnas.2005456117.

11. M. Bolles, D. Deming, K. Long, S. Agnihothram, A double-inactivated severe acute respiratory syndrome coronavirus vaccine provides incomplete protection in mice and induces increased eosinophilic …. Journal of (2011) (available at https://jvi.asm.org/content/85/23/12201.short).

12. D. Ricke, R. W. Malone, Medical Countermeasures Analysis of 2019-nCoV and Vaccine Risks for Antibody-Dependent Enhancement (ADE) (2020),, doi:10.2139/ssrn.3546070.

13. J. Wang, K. Tang, K. Feng, W. Lv, High Temperature and High Humidity Reduce the Transmission of COVID-19. SSRN Electronic Journal,, doi:10.2139/ssrn.3551767.

14. info coronavirus covid 19 - carte et donnees covid 19 en france. Gouvernement.fr, (available at https://www.gouvernement.fr/info-coronavirus/carte-et-donnees).

15. Field Briefing: Diamond Princess COVID-19 Cases, 20 Feb Update (2020), (available at https://www.niid.go.jp/niid/en/2019-ncov-e/9417-covid-dp-fe-02.html).

16. 新型コロナウイルス感染症について, (available at https://www.mhlw.go.jp/stf/seisakunitsuite/bunya/0000164708_00001.html).

17. N. M. Linton, T. Kobayashi, Y. Yang, K. Hayashi, A. R. Akhmetzhanov, S.-M. Jung, B. Yuan,R. Kinoshita, H. Nishiura, Incubation Period and Other Epidemiological Characteristics of 2019 Novel Coronavirus Infections with Right Truncation: A Statistical Analysis of Publicly Available Case Data. Journal of Clinical Medicine. 9 (2020), p. 538.

18. H. Nishiura, D. Klinkenberg, M. Roberts, J. A. P. Heesterbeek, Early epidemiological assessment of the virulence of emerging infectious diseases: a case study of an influenza pandemic. PLoS One. 4, e6852 (2009).

19. Stan Development Team, RStan: the R interface to Stan (2020), (available at http://mc-stan.org/).

20. Zhanwei Du, Xiaoke Xu, Ye Wu, Lin Wang, Benjamin J. Cowling, Lauren Ancel Meyers, Serial Interval of COVID-19 among Publicly Reported Confirmed Cases. Emerging Infectious Disease journal. 26 (2020), doi:10.3201/eid2606.200357.

21. Q. Bi, Y. Wu, S. Mei, C. Ye, X. Zou, Z. Zhang, X. Liu, L. Wei, S. A. Truelove, T. Zhang, W. Gao,C. Cheng, X. Tang, X. Wu, Y. Wu, B. Sun, S. Huang, Y. Sun, J. Zhang, T. Ma, J. Lessler, T. Feng, Epidemiology and Transmission of COVID-19 in Shenzhen China: Analysis of 391 cases and 1,286 of their close contacts. medRxiv, 2020.03.03.20028423 (2020).

22. L. Tindale, M. Coombe, J. E. Stockdale, E. Garlock, W. Y. V. Lau, M. Saraswat, Y.-H. B. Lee,L. Zhang, D. Chen, J. Wallinga, C. Colijn, Transmission interval estimates suggest pre-symptomatic spread of COVID-19. Epidemiology (2020),, doi:10.1101/2020.03.03.20029983.

23. S. Funk, socialmixr (Github; https://github.com/sbfnk/socialmixr).

24. O. Diekmann, J. A. Heesterbeek, J. A. Metz, On the definition and the computation of the basic reproduction ratio R0 in models for infectious diseases in heterogeneous populations. J. Math. Biol. 28, 365–382 (1990).

25. N. Hens, G. M. Ayele, N. Goeyvaerts, M. Aerts, J. Mossong, J. W. Edmunds, P. Beutels, Estimating the impact of school closure on social mixing behaviour and the transmission of close contact infections in eight European countries. BMC Infect. Dis. 9, 187 (2009).

26. Décret n° 2020-260 du 16 mars 2020 portant réglementation des déplacements dans le cadre de la lutte contre la propagation du virus covid-19 |Legifrance, (available at https://www.legifrance.gouv.fr/affichTexte.do?cidTexte=JORFTEXT000041728476&categorieLien=id).

27. C. I. Jarvis, K. Van Zandvoort, A. Gimma, K. Prem, CMMID COVID-19 working group, P. Klepac, G. J. Rubin, W. J. Edmunds, Quantifying the impact of physical distance measures on the transmission of COVID-19 in the UK. Epidemiology (2020),, doi:10.1101/2020.03.31.20049023.

28. J. Wallinga, M. Lipsitch, How generation intervals shape the relationship between growth rates and reproductive numbers. Proc. Biol. Sci. 274, 599–604 (2007).

